# Administration approaches of nursing assistants in hospitals: a scoping review

**DOI:** 10.1101/2022.03.20.22272663

**Authors:** Ben-tuo Zeng, Ying-hui Jin, Shu-dong Cheng, Yan-ming Ding, Ji-wei Du

## Abstract

**Objectives:** The administration of nursing assistants is closely associated with patient outcomes, but the current situation needs improvement. Studies evaluating intrahospital administration of nursing assistants are limited, and there is a lack of available evidence-based reviews in this field. This study aimed to identify existing literature on intrahospital nursing assistants’ administration approaches.

**Design:** Scoping review.

**Search strategy:** We searched fifteen electronic databases for English and Chinese articles published between January 2011 and March 2022. Publications on administration approaches, models and appraisal tools of intrahospital nursing assistants were included.

**Results:** Thirty-six studies were eligible for the review with acceptable quality. We identified one administration model, nine administration methods, fifteen educational programs, and seven appraisal tools from the included studies. The frequency effect size analysis yielded 15 topics of the main focus at four levels, suggesting that included articles were mainly (33%) focused on the competency of nursing assistants, and the lectures were the most (80%) used strategy in quality improvement projects. Evidence from the studies was of low-to-moderate quality, indicating huge gaps between evidence-based research and management practice.

**Conclusions:** A series of practical intrahospital administration approaches was revealed, and fifteen primarily focused topics were identified. We should explore this area more thoroughly using structured frameworks and standardized methodology. This scoping review will help managers find more effective ways to improve the quality of care. Researchers may focus more on evidence-based practice in nursing assistant administration using the 15 topics as a breakthrough.

**Strengths and limitations:**

1. First scoping review of practical administration approaches for nursing assistants in hospitals.
2. Presenting the main topics and focus of related articles.
3. Development of the nursing assistant administration was widely varied among countries.
4. Most of the included studies were of moderate-to-low methodological quality, and a huge gap exists between evidence-based research and management practice.

## BACKGROUND

Nursing assistants (NAs) are trained allied nursing personnel who provide or assist with basic care or support under the direction of onsite licensed nursing staff.^1^ In 2019, there were approximately 1.73 million NAs in the US, and this number in the European Union was 4.67 million in 2018,^2 3^ indicating that NAs have become the mainstay of care. As an integral part of routine healthcare, NAs are providing more care in hospitals due to the increasing aging population and the shortage of registered nurses (RNs) in recent years,^4 5^ thereby raising the criticality of nursing assistant administration. Nursing administration is now challenged by diverse educational backgrounds and the communication and connection between NAs and RNs.^6-8^

Standards and regulations of NAs have been established in several countries, e.g., OBRA 1987 Act in the US,^9^ the Cavendish Review and other documents in the European Union,^10 11^ and the Care Certificate in the UK.^12^ And in China, the National Health Commission emphasized the standardized regulation of NAs in 2019, providing instructions on NA training.^13^ However, the documents provided requirements for NA education, qualifications and training, but not for broader administration settings. There have not been uniform intrahospital NA administration regulations across countries.

The efficacy of the miscellaneous administration approaches followed by healthcare facilities was vague, leading to the grim current circumstances of NA administration. Lack of crucial competency, high turnover rate, low vocational identity, and low self-efficiency confused NAs and their administrators.^14-19^ The faultiness of these aspects would directly affect the quality of care. A higher turnover rate was associated with fewer infection events, and the retention rate was positively linked with clinical outcomes.^20 21^ The self-efficacy of NAs was associated with their burnout rate, which was very dangerous for care outcomes.^22^ In addition, communication among NAs, other nursing staff and administrators also needed improvement. NAs reported discontentment with administrators’ not understanding their work problems,^23^ and the performance of NAs also was influenced by relationships among NAs, RNs and other clinical staff.^24^

Various definitions, duties and unbalanced development of NAs worldwide^25 26^ have created barriers to identifying appropriate management strategies. In the US, NAs are also named certified nursing assistants (CNAs) and unlicensed assistive personnel (UAPs), and the names were nurse/nursing aides (NAs), health care assistants (HCAs) and nursing auxiliaries in the UK. Nursing attending workers (NAW) and nursing assistants (NA) were used in China.^26^ The communication of research from different countries is always confusing, and valid approaches and practices are needed to address the current chaos.

NAs take up several care duties that directly affect the outcomes of patients in hospitals. However, much of the previous research on nursing administration was developed in long-term care settings, and limited studies have been conducted in hospitals. Due to the scarcity of primary studies, organizing systematic reviews or other evidence syntheses is challenging, and therefore scoping reviews are needed and timely to identify existing evidence and assess the feasibility of conducting evidence-based research. In this article, we reviewed available administration approaches and assessment tools for NAs in inpatient care settings, described the current progress of this field, and presented a vision for further research and evidence synthesis.

## Aims and research questions

This review aimed to identify, describe and synthesize current knowledge and the existing literature on nursing assistants’ administration approaches and models, as well as education, skill training and multidimensional appraisal in hospitals. Two research questions were raised for intrahospital NA administration:

- What are the available approaches, models, programs, and tools?
- What are the most focused topics and most used methods of the existing studies?

## METHODS

### Study Design and Protocol

The review was conducted according to Joanna Briggs Institute (JBI) guidance and methodology for scoping review,^28 29^ and reported using the Preferred Reporting Items for Systematic reviews and Meta-analyses – Extension for Scoping Reviews (PRISMA-ScR).^27^ A structured protocol^30^ was prepared a priori according to the PRISMA Protocols (PRISMA-P) 2015 statement and explanation^31 32^ and PRISMA-ScR.

### Eligibility Criteria

This review was designed to identify studies which mainly focused on NAs, and discussed at least one administration-related topic. The inclusion criteria were:

1. Participants: nursing assistants (or certificated nursing assistants, nurse aides, etc.);
2. Setting: hospital;
3. Study type: Qualitative, quantitative, or mixed researches, evidence-based reviews, guidelines, consensus, or related dissertations;
4. Focus: specific approaches or models, or overall administration models, programs, tools or frameworks;
5. Methodology: reported clear intervention/exposure methods and evaluation tools;
6. Language: English or Chinese;
7. Peer-reviewed studies published between January 2011 and March 2022.

The exclusion criteria were: (1) Participants: other healthcare personnel, students, or orderlies; or mixed participants with different occupations, and nursing assistants were not discussed or presented separately; (2) Focus: focusing on specific areas, and the conclusions were only fit for the focused areas. (3) Setting: long-term care facilities, nursing homes, or skilled nursing facilities; and (4) Outcome: not reported key administrational outcomes.

### Search Strategy and Study Selection

For initial screening, we retrieved comprehensive, medical, nursing and evidence-based databases as follows: medical databases (PubMed, Embase, APA PsycInfo, Wanfang Med and SinoMed); nursing databases (CINAHL and Ovid Emcare), general databases (Scopus, ProQuest and CNKI) and evidence-based practice databases (NICE, AHRQ, CADTH, JBI EBP and Cochrane DSR). The initial search was completed in November 2021, and we updated the results in March 2022.

We ran a preliminary search in PubMed to identify keywords, search fields, and related topics. We used PubMed PubReMiner^33^ to identify related keywords for search strategy establishment. Afterward, search strategies in each database were developed, including key terms: nursing assistants, nursing aides, nursing auxiliar*, administr*, educat*, training, apprais*, organization and administration. Full search strategy is displayed in **online supplemental file 1**. References of all included studies were manually searched using terms “assistant” and “aide”.

All publications were imported into EndNote 20.2 (build 15709) for citation management, and duplicates were removed. A brief screening checklist (**online supplemental file 2**) was developed for study selection to minimize the inconsistency of the reviewers. Two reviewers screened all the studies independently according to the eligibility criteria. Divergences were discussed by the reviewers together or with a third researcher.

### Data Charting

Article characteristics, sample size and participant demographics, focused topics, study designs and outcomes were charted from eligible studies. Two researchers designed a structured data charting tool (online supplemental file 3) for data charting and continuously refined it. Data charting of all eligible studies was performed by two authors independently and was corroborated by a third researcher.

### Quality Appraisal

The Mixed Methods Appraisal Tool (MMAT) Version 2018^34 35^ was applied for qualitative, quantitative or mixed-methodological studies. AMSTAR 2, a critical appraisal tool for systematic reviews^36^, was used for the critical appraisal of evidence-based reviews. An overall score was carried out for each included study. For the MMAT, we calculated the percentage of items answered “Yes” in Section 2, and for AMSTAR 2, after items that were not applicable were excluded, we conducted a grade of overall confidence according to the criteria by Shea et al.^36^

### Data Synthesis

Eligible studies were divided into administration approaches, education and training, and appraisal tools. We summarized the study types, main focus and detailed intervention/exposure measures for the administration, education and training fields. For studies on practical tools, detailed tool information and psychometrics were extracted. For further interpretation, we conducted a frequency effect size analysis of the area of the main focuses and intervention strategies based on the calculating effect size method from the metasummary methodology introduced by Sandelowski et al.^37^

### Patient and Public Involvement Statement

It was not appropriate or possible to involve patients or the public in the design, or conduct, or reporting, or dissemination plans of our research.

## RESULTS

The search identified 1,973 related studies, among which 538 were Chinese publications. 138 publications remained for full-text screening, where 103 articles were excluded (see **online supplemental file 4**). Thirty-five studies from databases were included in the scoping review, and one study published from 2011 to 2021 were manually identified from the reference lists. Ultimately, we identified 36 eligible studies^38-73^ for our scoping review (**figure 1**).

**Figure 1.**
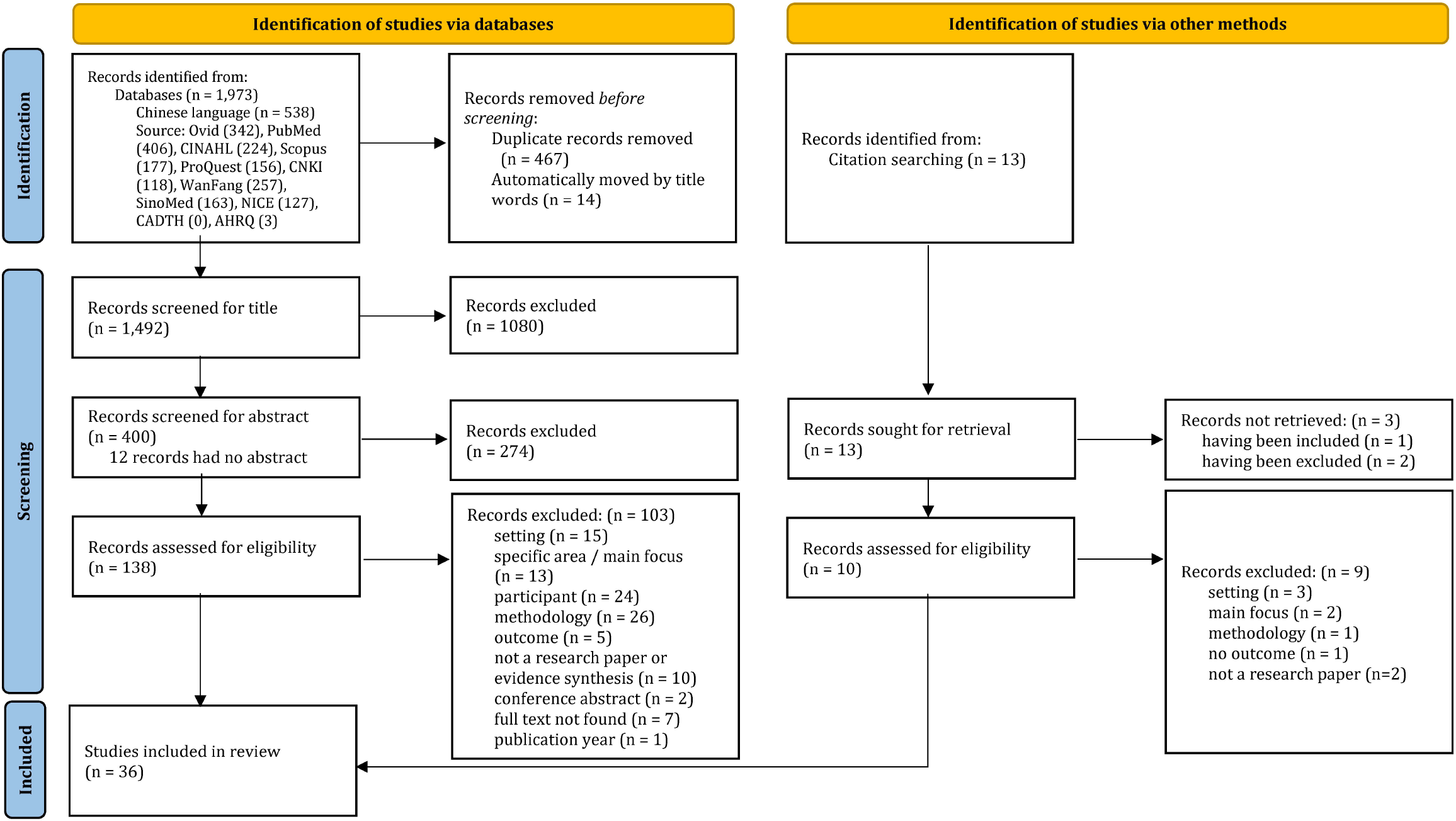
PRISMA flow diagram for study selection

### Study Characteristics

Study characteristics and contents are displayed in **table 1**. Settings, limitations, fundings and competing interests of the studies are shown in **online supplemental file 5**.

**Table 1.**
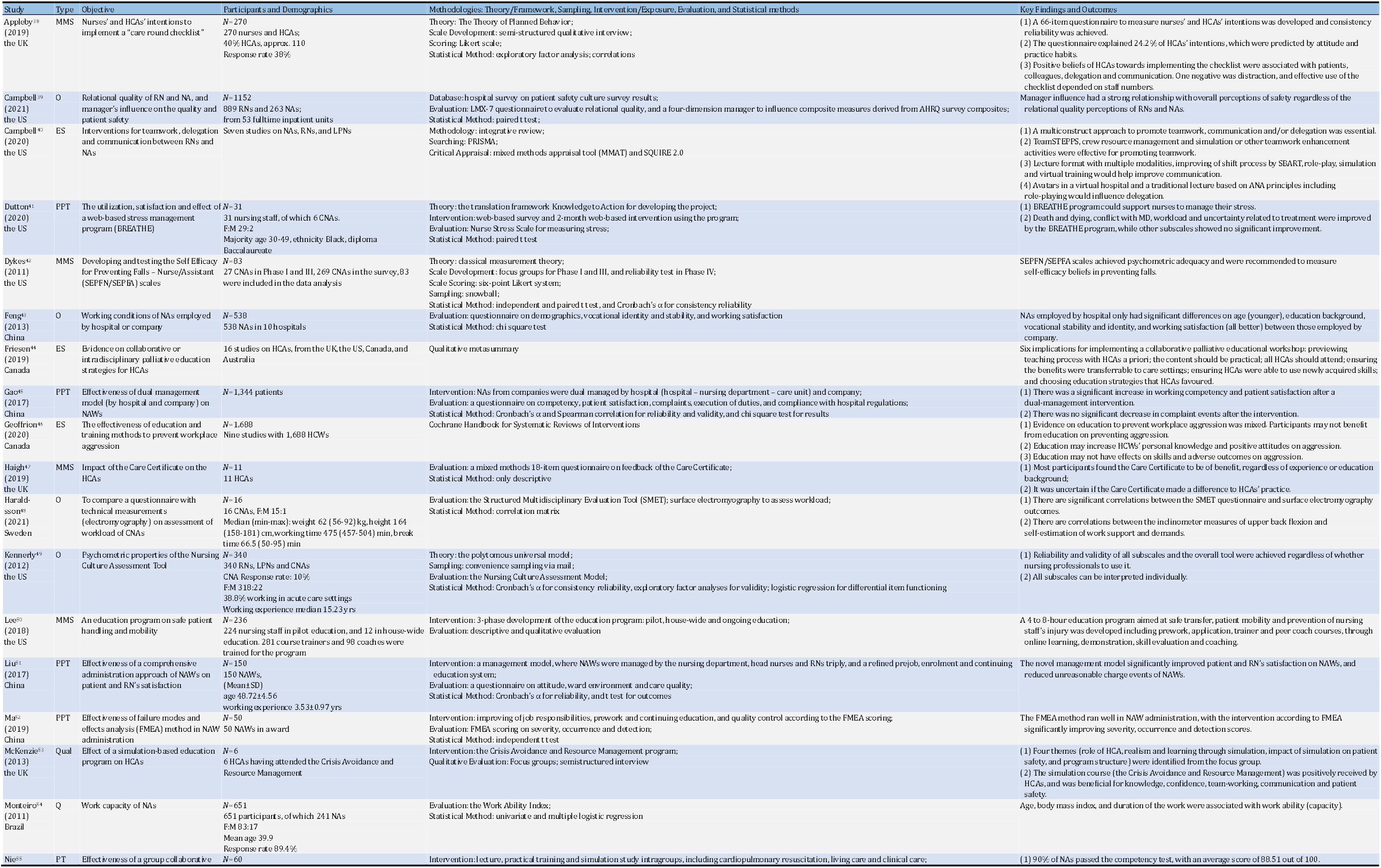
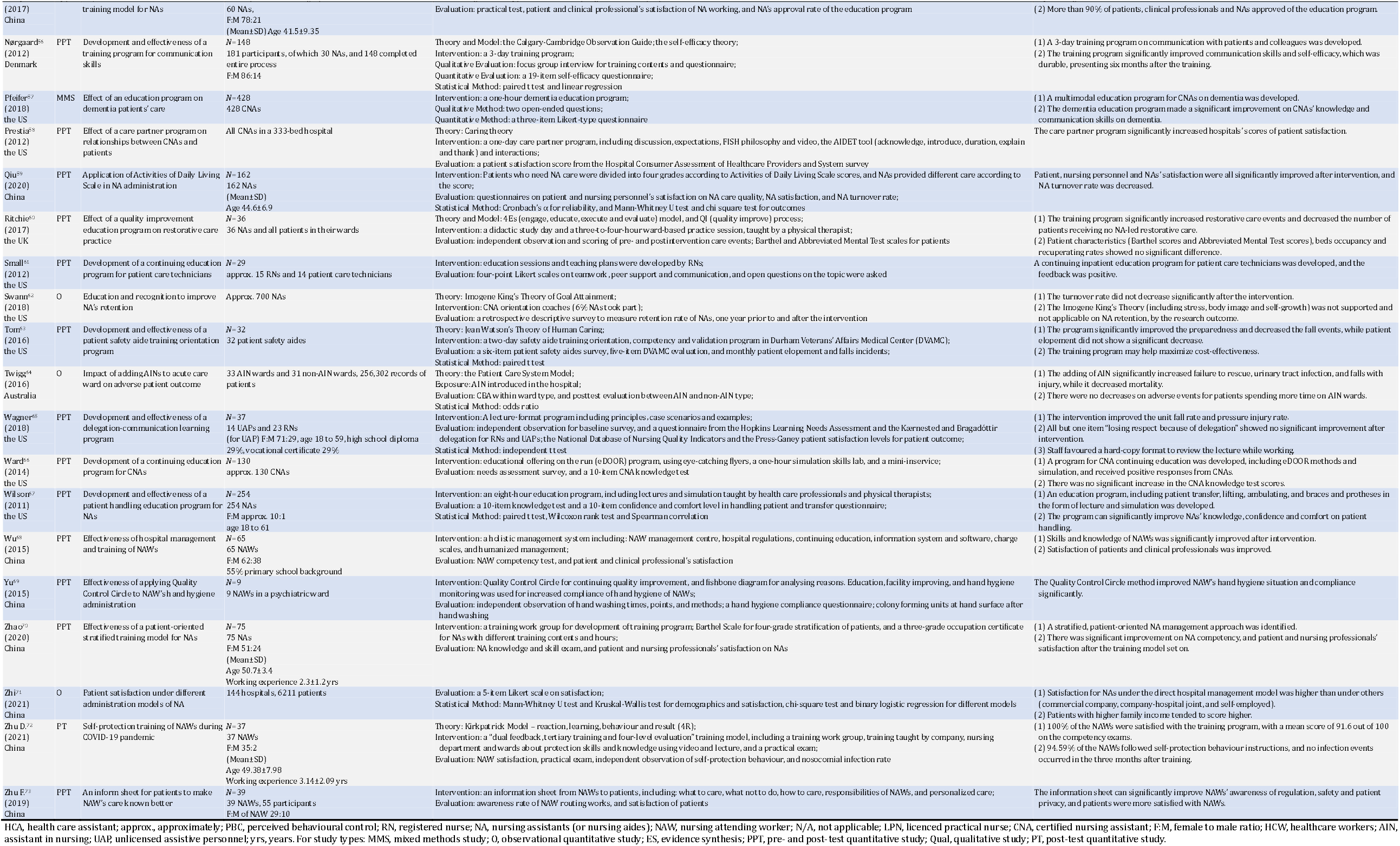
Study Characteristics

The articles were mainly from the US and China, and 22 of 36 studies were published in the last five years (2017-2022). Two theses, three evidence syntheses and 31 original research papers were included. 25 articles presented quantitative designs (17 interventional and eight descriptive), one was qualitative, and seven studies employed mixed methods. Most studies (19 of 24) with interventions applied a quasi-experimental, pre- and post-test design, while one study^57^ presented a retrospective post-then-pretest design.^74^ Focus group interviews were the most employed method in qualitative and mixed methods studies. The three evidence syntheses were in different types: one integrated review,^40^ one qualitative metasummary,^44^ and one systematic review.^46^

All 33 original studies were conducted in hospitals due to our inclusion criteria, and the sample size ranged from six to 700 NAs. Three studies^45 64 71^ were focused on patients’ attitudes, and did not report the demographics of NAs. Only one study^54^ reported a higher than 60% response rate, while others ranged from 10% to 40%.

The methodological quality appraisal results of the studies are listed in **online supplemental file 6**. The mean score of all 33 original studies assessed by the MMAT reached a level of 65%, and for evidence-based reviews, two of the studies^40 44^ received a “very low” rating, while the other one^46^ got a “high”. Overall, the methodological quality of the included studies was considered acceptable.

### Approaches and Models

Fourteen studies addressed various administration approaches and focuses, as listed in **table 2**. The development of NA administration in China was still preliminary, and articles by Chinese researchers were more fixated on employment models. Five studies^43 45 51 68 71^ were aimed at the change of management and employment from company-led to hospital-led, or double-track, with consistently positive results on satisfaction and NA/NAW working competency after intervention. Other original studies identified four practical tools or methods, i.e., the Failure Mode and Effects Analysis (FMEA),^52^ the Activities of Daily Living (ADL) scale,^59^ the Quality Control Circle,^69^ and an information sheet to patients,^73^ as well as two programs on stress and CNA-patient relationships.^41 58^

**Table 2.**
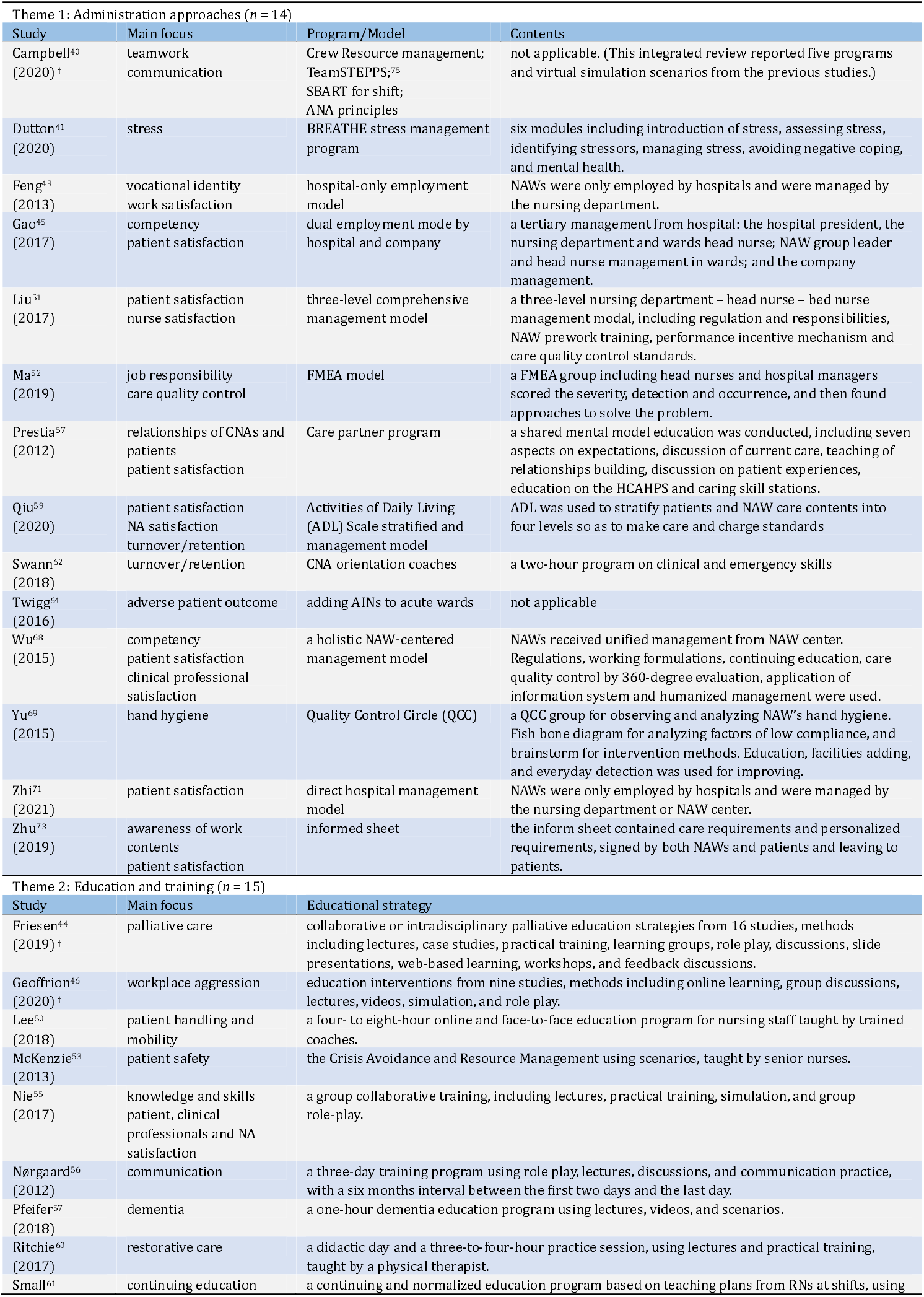
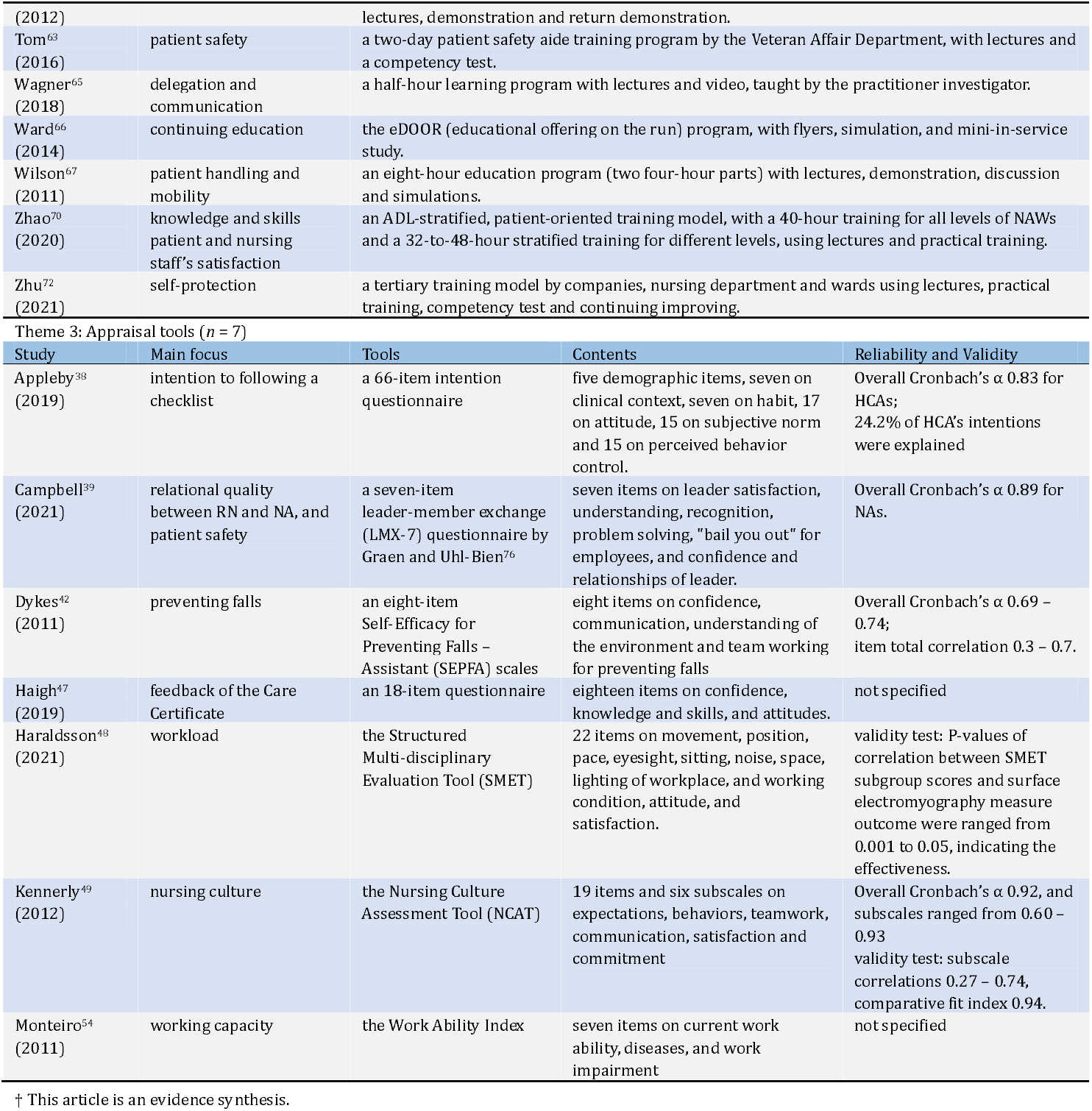
Administration approaches, training & education programs, appraisal tools and main focuses (*n* = 36)

| Theme 1: Administration approaches (*n* = 14) |  |  |  |
| --- | --- | --- | --- |
| Study | Main focus | Program/Model | Contents |
| Campbell<sup>40</sup><br>(2020) † | teamwork<br>communication | Crew Resource management;<br>TeamSTEPPS;<sup>75</sup><br>SBART for shift;<br>ANA principles | not applicable. (This integrated review reported five programs and virtual simulation scenarios from the previous studies.) |
| Dutton<sup>41</sup><br>(2020) | stress | BREATHE stress management program | six modules including introduction of stress, assessing stress, identifying stressors, managing stress, avoiding negative coping, and mental health. |
| Feng<sup>43</sup><br>(2013) | vocational identity<br>work satisfaction | hospital-only employment model | NAWs were only employed by hospitals and were managed by the nursing department. |
| Gao<sup>45</sup><br>(2017) | competency<br>patient satisfaction | dual employment mode by hospital and company | a tertiary management from hospital: the hospital president, the nursing department and wards head nurse; NAW group leader and head nurse management in wards; and the company management. |
| Liu<sup>51</sup><br>(2017) | patient satisfaction<br>nurse satisfaction | three-level comprehensive management model | a three-level nursing department – head nurse – bed nurse management modal, including regulation and responsibilities, NAW prework training, performance incentive mechanism and care quality control standards. |
| Ma<sup>52</sup><br>(2019) | job responsibility<br>care quality control | FMEA model | a FMEA group including head nurses and hospital managers scored the severity, detection and occurrence, and then found approaches to solve the problem. |
| Prestia<sup>57</sup><br>(2012) | relationships of CNAs and patients<br>patient satisfaction | Care partner program | a shared mental model education was conducted, including seven aspects on expectations, discussion of current care, teaching of relationships building, discussion on patient experiences, education on the HCAHPS and caring skill stations. |
| Qiu<sup>59</sup><br>(2020) | patient satisfaction<br>NA satisfaction<br>turnover/retention | Activities of Daily Living (ADL) Scale stratified and management model | ADL was used to stratify patients and NAW care contents into four levels so as to make care and charge standards |
| Swann<sup>62</sup><br>(2018) | turnover/retention | CNA orientation coaches | a two-hour program on clinical and emergency skills |
| Twigg<sup>64</sup><br>(2016) | adverse patient outcome | adding AINs to acute wards | not applicable |
| Wu<sup>68</sup><br>(2015) | competency<br>patient satisfaction<br>clinical professional satisfaction | a holistic NAW-centered management model | NAWs received unified management from NAW center. Regulations, working formulations, continuing education, care quality control by 360-degree evaluation, application of information system and humanized management were used. |
| Yu<sup>69</sup><br>(2015) | hand hygiene | Quality Control Circle (QCC) | a QCC group for observing and analyzing NAW's hand hygiene. Fish bone diagram for analyzing factors of low compliance, and brainstorm for intervention methods. Education, facilities adding, and everyday detection was used for improving. |
| Zhi<sup>71</sup><br>(2021) | patient satisfaction | direct hospital management model | NAWs were only employed by hospitals and were managed by the nursing department or NAW center. |
| Zhu<sup>73</sup><br>(2019) | awareness of work contents<br>patient satisfaction | informed sheet | the inform sheet contained care requirements and personalized requirements, signed by both NAWs and patients and leaving to patients. |
| Theme 2: Education and training (*n* = 15) |  |  |  |
| Study | Main focus | Educational strategy |  |
| Friesen<sup>44</sup><br>(2019) † | palliative care | collaborative or interdisciplinary palliative education strategies from 16 studies, methods including lectures, case studies, practical training, learning groups, role play, discussions, slide presentations, web-based learning, workshops, and feedback discussions. |  |
| Geoffrion<sup>46</sup><br>(2020) † | workplace aggression | education interventions from nine studies, methods including online learning, group discussions, lectures, videos, simulation, and role play. |  |
| Lee<sup>50</sup><br>(2018) | patient handling and mobility | a four- to eight-hour online and face-to-face education program for nursing staff taught by trained coaches. |  |
| McKenzie<sup>53</sup><br>(2013) | patient safety | the Crisis Avoidance and Resource Management using scenarios, taught by senior nurses. |  |
| Nie<sup>55</sup><br>(2017) | knowledge and skills<br>patient, clinical professionals and NA satisfaction | a group collaborative training, including lectures, practical training, simulation, and group role-play. |  |
| Nørgaard<sup>56</sup><br>(2012) | communication | a three-day training program using role play, lectures, discussions, and communication practice, with a six months interval between the first two days and the last day. |  |
| Pfeifer<sup>57</sup><br>(2018) | dementia | a one-hour dementia education program using lectures, videos, and scenarios. |  |
| Ritchie<sup>60</sup><br>(2017) | restorative care | a didactic day and a three-to-four-hour practice session, using lectures and practical training, taught by a physical therapist. |  |
| Small<sup>61</sup> | continuing education | a continuing and normalized education program based on teaching plans from RNs at shifts, using |  |

|  |  |  |
| --- | --- | --- |
| (2012) |  | lectures, demonstration and return demonstration. |
| Tom<sup>63</sup><br>(2016) | patient safety | a two-day patient safety aide training program by the Veteran Affairs Department, with lectures and a competency test. |
| Wagner<sup>65</sup><br>(2018) | delegation and communication | a half-hour learning program with lectures and video, taught by the practitioner investigator. |
| Ward<sup>66</sup><br>(2014) | continuing education | the eDOOR (educational offering on the run) program, with flyers, simulation, and mini-in-service study. |
| Wilson<sup>67</sup><br>(2011) | patient handling and mobility | an eight-hour education program (two four-hour parts) with lectures, demonstration, discussion and simulations. |
| Zhao<sup>70</sup><br>(2020) | knowledge and skills<br>patient and nursing staff's satisfaction | an ADL-stratified, patient-oriented training model, with a 40-hour training for all levels of NAWs and a 32-to-48-hour stratified training for different levels, using lectures and practical training. |
| Zhu<sup>72</sup><br>(2021) | self-protection | a tertiary training model by companies, nursing department and wards using lectures, practical training, competency test and continuing improving. |

Theme 3: Appraisal tools (n = 7)
| Study | Main focus | Tools | Contents | Reliability and Validity |
| --- | --- | --- | --- | --- |
| Appleby<sup>38</sup><br>(2019) | intention to following a checklist | a 66-item intention questionnaire | five demographic items, seven on clinical context, seven on habit, 17 on attitude, 15 on subjective norm and 15 on perceived behavior control. | Overall Cronbach's $\alpha$ 0.83 for HCAs; 24.2% of HCA's intentions were explained |
| Campbell<sup>39</sup><br>(2021) | relational quality between RN and NA, and patient safety | a seven-item leader-member exchange (LMX-7) questionnaire by Graen and Uhl-Bien<sup>76</sup> | seven items on leader satisfaction, understanding, recognition, problem solving, "bail you out" for employees, and confidence and relationships of leader. | Overall Cronbach's $\alpha$ 0.89 for NAs. |
| Dykes<sup>42</sup><br>(2011) | preventing falls | an eight-item Self-Efficacy for Preventing Falls – Assistant (SEPFA) scales | eight items on confidence, communication, understanding of the environment and team working for preventing falls | Overall Cronbach's $\alpha$ 0.69 – 0.74; item total correlation 0.3 – 0.7. |
| Haigh<sup>47</sup><br>(2019) | feedback of the Care Certificate | an 18-item questionnaire | eighteen items on confidence, knowledge and skills, and attitudes. | not specified |
| Haraldsson<sup>48</sup><br>(2021) | workload | the Structured Multi-disciplinary Evaluation Tool (SMET) | 22 items on movement, position, pace, eyesight, sitting, noise, space, lighting of workplace, and working condition, attitude, and satisfaction. | validity test: P-values of correlation between SMET subgroup scores and surface electromyography measure outcome were ranged from 0.001 to 0.05, indicating the effectiveness. |
| Kennerly<sup>49</sup><br>(2012) | nursing culture | the Nursing Culture Assessment Tool (NCAT) | 19 items and six subscales on expectations, behaviors, teamwork, communication, satisfaction and commitment | Overall Cronbach's $\alpha$ 0.92, and subscales ranged from 0.60 – 0.93<br>validity test: subscale correlations 0.27 – 0.74, comparative fit index 0.94. |
| Monteiro<sup>54</sup><br>(2011) | working capacity | the Work Ability Index | seven items on current work ability, diseases, and work impairment | not specified |

† This article is an evidence synthesis.

Negative outcomes were found in turnover rates after a CNA orientation coach^62^ and in adverse events with the addition of AINs to acute wards.^64^ Meanwhile, several approaches, including crew resource management, the TeamSTEPPS program,^75^ and the SBART shift model, were summarized by an integrated review from Campbell et al.^40^

### Education and Training Programs

Each of the 13 research papers developed an education program with different topics, while two evidence reviews^44 46^ discussed educational strategies on palliative care and workplace aggression with wide availability (table 2). NA knowledge and skills were most emphasized. Six studies highlighted five aspects of NA competency: patient handling,^50 67^ palliative care,^44^ dementia,^57^ restorative care^60^ and routine work capacity.^55^ Of other studies, two^53 63^ improved patient safety, two^56 65^ were on communication, two^61 66^ pointed to continuing education and the other two^46 72^ focused on self-protection. The strategies taken ranged from classical lectures to web-based learning, simulation and practical training. The mixed outcome of education on workplace aggression was reviewed by Geoffrion et al.,^46^ addressing the need of further study in this area.

### Appraisal Tools

We found seven valid appraisal tools for NA administration with stable psychometrics from the included studies (table 2):

1. HCA & RNs’ intention questionnaire;^38^
2. LMX-7 relational quality questionnaire from Campbell et al.^39^and developed by Graen et al.;^76^
3. NA working questionnaire^47^;
4. the Nursing Culture Assessment Tool (NCAT);^49^
5. the Structured Multidisciplinary Evaluation Tool (SMET) from Haraldsson et al.^48^ and developed by the author in 2016;^77^
6. Self-Efficacy for Preventing Falls – Assistants (SEPFA);^42^
7. the Work Ability Index from Monteiro et al.^54^ and produced by Ilmarinen.^78^

Three of seven papers developed original scales and tested reliability and validity. Four studies examined the psychometrics of existing questionnaires or applied questionnaires to NA administration and reported eligible outcomes. The main focuses ranged from working capacity and workload to relationships, intentions and nursing culture.

### Frequency Effect Size Analysis

We conducted frequency effect size analysis on the included studies’ main focuses and strategies (**table 3** and **figure S1** in **online supplemental file 7**). Fifteen main focuses were identified at four levels. The most reported focuses were competency at the NA level (frequency effect size 33%), communication and clinical staff satisfaction (both 14%) at the clinical personnel level, and patient satisfaction (25%) at the patient level. Both topics at the fourth facility management level (retention and care quality control) were at a 6% level of effect size.

**Table 3.** Frequency effect size analysis of main focuses and intervention methods

| Topics | Studies | Frequency effect size |
| --- | --- | --- |
| Main focus (n = 36) |  |  |
| Level 1: NA |  |  |
| competency (skills and knowledge) | 44, 45, 47, 50, 52, 54, 55, 57, 60, 67, 68, 70 | 33% |
| self-efficacy, job-related identity and responsibility | 43, 47, 49, 52, 73 | 14% |
| satisfaction | 43, 49, 52, 55, 59 | 14% |
| continuing education | 61, 66, 68 | 8% |
| self-protection | 46, 72 | 6% |
| workload and stress | 41, 48 | 6% |
| intention | 38 | 3% |
| Level 2: clinical personnel |  |  |
| communication and teamwork | 40, 47, 49, 56 | 14% |
| clinical/nursing staff satisfaction | 51, 55, 59, 68, 70 | 14% |
| relationship | 39 | 3% |
| Level 3: patient |  |  |
| patient satisfaction | 45, 51, 55, 58, 59, 68, 70, 71, 73 | 25% |
| patient safety | 42, 45, 53, 63, 64 | 11% |
| relationships | 58 | 3% |
| Level 4: facility management |  |  |
| retention/turnover | 59, 62 | 6% |
| care quality control | 52, 69 | 6% |
| Intervention method |  |  |
| Education & training (n = 20) |  |  |
| lecture | 50-52, 55, 56-58, 60, 61, 63, 65, 67-70, 72 | 80% |
| simulation/role play | 50, 55, 58, 66-68 | 30% |
| practical training | 52, 55, 56, 68, 70, 72 | 30% |
| exam | 62, 63, 66, 69, 72 | 25% |
| discussion | 50, 56, 58, 67 | 20% |
| video | 50, 57, 65, 68 | 20% |
| demonstration | 61, 67-69 | 20% |
| web-based training | 41, 50 | 10% |
| group study | 50, 55 | 10% |
| scenario | 53, 57 | 10% |
| flyer | 66 | 5% |

Twenty studies contained the theme of education and training strategies. Face-to-face lectures were still the most employed method (80%), followed by simulation, role play, and practical training (30%). We also noted that flyers with various knowledge and skills for CNA continuing education were designed by Ward et al.,^66^ which was a unique and effective method not adopted by other studies. Due to the significant heterogeneity, we failed to distinguish themes in studies on NA administration methods and only summarized their objectives. Approximately 62% of the studies evaluated tools for administration, and the other five papers were on hospital management models (38%).

## DISCUSSION

This review outlined existing administration tools, management models, education programs and appraisal scales from previous studies. The results implied a need to investigate more on administration models and frameworks for NA administration in hospitals. High-quality evidence of the efficacy of existing educational strategies was scarce. Gaps in NA administration development and current circumstances between developed and developing areas were noted and needed improvement.

### Approaches and Models

Existing articles for administration approaches, models, and frameworks were scarce. Only fourteen studies presented evaluations on these topics, and the focuses were region-specific, i.e., researchers from China and the Europe-US region typically targeted different objectives.

In China, NAWs were employed by companies or hospitals, or were self-employed.^79 80^ The company employment model took the majority.^81^ The selection and training of NAWs were conducted by directors and executors of companies but not clinical professionals, leading to uncontrolled quality of care and muddled management.^79 82^ The researchers proposed to improve this context. Included Chinese studies showed a three-tiered hierarchical model where NAWs were managed by (1) nurse departments or NAW centers, (2) head nurses and (3) ward nurses. However, this review revealed remarkable heterogeneity and unrepresentative sampling in this area. Further investigation of the hospital management model and the effectiveness of other mature models in Chinese hospitals can be the next step.

Chinese researchers also evaluated the FMEA model, ADL scale and Quality Control Circle (QCC). FMEA and QCC were widely-used mature models in hospital routine management^83-86^, but have rarely been evaluated for NAs. However, without any new points added, the ideas were only a panning from nurses to NAs. On the other hand, the application of ADL scale was more innovative. ADL was first developed by Katz^87^ and is one of the most widely used tools for assessing patient functions. Different ADL scores and levels represented patients’ statuses and care needs to guide managers to a more cost-effective and patient-oriented model of NA allocation.

For researchers from the countries where NA industry has been highly developed, a holistic program was typically their focus. They also focused on more humanistic aspects such as communication, workload, safety, etc. They^41 58^ developed or evaluated programs to improve administration, i.e., the TeamSTEPPS program, BREATHE program and the Care Partner Programs. The programs were well-designed and of good availability, but the small sample size still placed barriers to their widespread application. Additional studies on approaches to widen the applicability and design of more diverse programs are crucial.

The difference in the hotspots of the research area was strongly related to the degree of development of NA industry in different regions. Our included studies were strongly demand-driven. In a region where NA regulations were not yet well-established, relevant studies tended to be more primary and discuss basic administration structures, e.g., employment modes. On the contrary, in countries where NAs were mandated by law to be employed and managed by healthcare institutes themselves, the topic was no longer necessary for research, and researchers have turned to explore more advanced knowledge.

### Educational Strategies

Eleven groups of educational strategies were applied to 20 included studies. 90% of the studies employed at least two groups of strategies, with an average of 2.60, and multiple education methods may result in more positive outcomes compared to single-method teaching.^88^ A simulation was highlighted because it has been shown to be effective in nursing education,^89 90^ and a debrief simulation method was recommended.^91^ Web-based training has become a modern trend, especially during the COVID-19 pandemic.^92^ However, the evidence regarding the effectiveness of e-learning in nursing education remains unclear.^93 94^ Moreover, half of the articles used interactive methods, e.g., simulation, discussion and group study. We found this to be a trend in NA education and training in recent years. When transferring these teaching methods to NAs, attention should be paid to the educational background gap between NAs and nursing students.

### Focuses and Topics

Twenty-three focuses were initially found by the frequency effect analysis, where 15 topics were distilled. NA knowledge and skills were most widely considered, with a trend of being more specified for improving competency. Most of the studies conducted an on-the-job training mode, thus the necessity of exploring advanced competency to avoid repeats. Focuses that contributed smaller effect sizes may denote the possible directions of future studies. NA intentions, relationships of NA-other nursing staff and NA-patients, workload, stress and retention in hospitals needed more investigation. Existing studies revealed that all of the factors above influenced the quality of care at the RN or nursing home level,^95-98^ but evidence of influence and improvement methods of NAs in hospitals is limited.

A noteworthy point was that much of the research in this area of NA was essentially a transposition of methods previously applied among RNs to NAs, within all three fields this review investigated. FMEA, QCC, and patient satisfaction were significant examples. However, as discussed above, researchers should be very careful when dealing with the differences in educational background and job content between RNs and NAs.

### Negative Outcomes

Three studies^46 62 64^ reported notable negative or mixed outcomes. The systematic review from Geoffrion^46^ et al. supposed that both patients and healthcare workers may not benefit from educational programs on workplace aggression for clinical staff, revealing that other approaches or the education for patients may be conducted for the improvement of NA safety. Swann^62^ evaluated the influence of the CNA orientation coaches on the retention rate and derived a negative result, while Twigg^64^ placed an analysis of adding AINs to acute care wards with unexpected outcomes on failure to rescue, urinary tract infection, and falls. The sample of the latter two articles was still limited, with the potential risk of inadequate study designs (observational study to evaluate the interventions), so researchers may conduct more studies on their topics in spite of the discouraging results.

### Gaps

Several gaps were concluded for the NA administration area: (1) NA definitions, regulations and circumstances varied widely among countries, especially between high-income and those with moderate to low incomes, which created barriers to global evidence and practical experience shared processes. (2) The limited sample sizes and nonrandomized study designs of the included studies may decrease the reliability of outcomes. (3) The included studies were of low-to-moderate quality of evidence and availability, and the study design was not reported in-detailed. There were barriers between existing studies and evidence-based practice. (4) Theories and conceptual frameworks were often neglected in the study designs.

### Limitations

The diversified intervention methods of the included studies led to significant heterogeneity, so barriers existed for further analysis and synthesis. We included only Chinese and English articles, while studies published in other languages were excluded, thus leading to a potential risk of bias. Furthermore, more studies may not be included for analysis in other social sciences databases, as NA administration is a broad, multidisciplinary and interdisciplinary topic. We also noted that three included evidence-based reviews addressed 32 original studies, where 13 studies were published between 2011 and 2022 but were not included in our scoping review, implying a potential risk of incomprehensiveness of our work.

## CONCLUSION

This scoping review demonstrated the practical administration approaches and focus from previous studies for hospital nursing assistants. The review found a total of nine administration methods, one administration model, 15 education and training programs, and seven appraisal tools. With the frequency effect size analysis, 15 main focus groups and 11 educational strategies used for improving administration were outlined. The insight from our review will add knowledge to effective NA administration for hospital managers and head nurses and help to improve the quality of care with increasing evidence.

Barriers remain between the intrahospital NA administration area and evidence-based nursing research and practice. The endeavour to apply evidence-based methods to administration will be arduous but will contribute greatly to improved outcomes.

We expect that the administration approaches concluded by our study will help leaders interpret more about effective management to improve quality of care and benefit all clinical staff and patients. The difference between hospitals and long-term care settings should be recognized, and more studies on NAs in hospitals are expected. Researchers should draw more attention to evidence-based methods in the administration area, resulting in continuous improvement, global sharing and system establishment of intrahospital nursing assistants’ administration.

## Data Availability

Data Availability Statement: The data that supports the findings of this study are available in the supplementary material of this article, and the protocol of this study are openly available in ResearchGate at http://doi.org/10.13140/RG.2.2.29106.12483/1

## Acknowledgements

We acknowledge Ms. Zhuo Lin and Ms. Zhang Yu-jing for their ideas on study design, data synthesis, and manuscript refinement.

## Contributors

ZB and CS performed study selection, data charting and quality appraisal. ZB and JY completed the study design, search, data synthesis and composition of the manuscript, and study design was also refined by Ms. Zhuo Lin and Ms. Zhang Yu-jing. Any raised disagreement was discussed and solved by ZB, CS, JY and DJ. All authors and Ms. Zhuo Lin, Ms. Zhang Yu-jing contributed to the revisions and refinement of the manuscript.

## Funding

The authors received no financial support for the research, authorship or publication of this article. Competing Interests: The authors declare that there is no conflict of interest.

## Ethics Approval

A scoping review does not contain human or animal-related objectives, tissues or data, and all data used in this study were from publicly available source, so it is not necessary for this review to have an ethical approval.

## Data Availability Statement

Data are available in a public, open access repository. The data that supports the findings of this study are available in the supplementary material of this article, and the protocol of this study are openly available (DOI: 10.13140/RG.2.2.29106.12483/1)

## Notes

### Competing Interest Statement

The authors have declared no competing interest.

### Clinical Protocols

http://doi.org/10.13140/RG.2.2.29106.12483/1

### Funding Statement

This study did not receive any funding.

### Summary of Updates

We revised this article after receiving comments from reviewers. Since the peer review process and the publication of this article require time, we revised this preprint to follow the latest version.

## References

1 U.S. Bureau of Labor Statistics. 2018 SOC Definitions. Available: https://www.bls.gov/soc/2018/soc_2018_definitions.pdf [Accessed 2 Dec 2021].

2 Eurostat. Healthcare personnel statistics - nursing and caring professionals. Available: https://ec.europa.eu/eurostat/statistics-explained/index.php?title=Healthcare_personnel_statistics_-_nursing_and_caring_professionals [Accessed 6 Nov 2021].

3 Jumabhoy S, Jung HY, Yu J. Characterizing the direct care health workforce in the United States, 2010-2019. Journal of American Geriatric Society 2022;70(2):512–21.

4 NSI Nursing Solutions. 2021 NSI National Health Care Retention & RN Staffing Report. Available: https://www.nsinursingsolutions.com/Documents/Library/NSI_National_Health_Care_Retention_Report.pdf [Accessed 19 Nov 2021]

5 Zhang X, Tai D, Pforsich H, et al. United States registered nurse workforce report card and shortage forecast: a revisit. Am J Med Qual 2017;33(3):229–36.

6 Blay N, Roche MA. A systematic review of activities undertaken by the unregulated nursing assistant. J Adv Nurs 2020;76(7):1538–51.

7 Campbell AR, Kennerly S, Swanson M, et al. Relational quality between the RN and nursing assistant: essential for teamwork and communication. J Nurs Adm 2021;51(9):461–7.

8 Office for National Statistics. SOC 2020 Volume 2: the coding index and coding rules and conventions. Available: https://www.ons.gov.uk/methodology/classificationsandstandards/standardoccupationalclassificationsoc/soc2020/soc2020volume2codingrulesandconventions [Accessed 20 Nov 2021].

9 Omnibus Budget Reconciliation Act of 1987: Conference report filed in House, H. Rept. 100-495, 100th Congress December (1987).

10 Cavendish C. The Cavendish Review. Available: https://assets.publishing.service.gov.uk/government/uploads/system/uploads/attachment_data/file/236212/Cavendish_Review.pdf [Accessed 13 Nov 2021].

11 Eurodiaconia. The education, training and qualifications of nursing and care assistants across Europe. Available: https://www.eurodiaconia.org/wordpress/wp-content/uploads/2016/08/The-education-training-and-qualifications-of-nursing-and-care-assistants-across-Europe-Final.pdf [Accessed 16 Nov 2021].

12 Skills for Care, Skills for Health, Health Education England. The Care Certificate Workbook. Available: https://www.skillsforcare.org.uk/Learning-development/inducting-staff/care-certificate/Care-Certificate-workbook.aspx [Accessed 16 Nov 2021].

13 Guojia weishengjiankang weiyuanhui [National Health Commision of the People’s Republic of China]. [Strengthening the training and standardized management of nursing assistants]. Available: http://www.nhc.gov.cn/cms-search/xxgk/getManuscriptXxgk.htm?id=f239ab4290f94d3cb6b36d1705e29f34 [Accessed 13 Nov 2021].

14 Burgio LD, Stevens A, Burgio KL, et al. Teaching and maintaining behavior management skills in the nursing home. Gerontologist 2002;42(4):487–96.

15 Centers for Disease Control and Prevention. National Nursing Assistant Survey. Available: https://www.cdc.gov/nchs/nnhs/nnas.htm [Accessed 19 Nov 2021].

16 Cready CM, Yeatts DE, Gosdin MM, et al. CNA empowerment: Effects on job performance and work attitudes. J Gerontol Nurs 2008;34(3):26–35.

17 Gion T, Abitz T. An approach to recruitment and retention of certified nursing assistants using innovation and collaboration. J Nurs Adm 2019;49(7/8):354–8.

18 Probst JC, Baek JD, Laditka SB. Characteristics and recruitment paths of certified nursing assistants in rural and urban nursing homes. J Rural Health 2009;25(3):267–75.

19 Temple A, Dobbs D, Andel R. Exploring correlates of turnover among nursing assistants in the National Nursing Home Survey. Health Care Manage Rev 2009;34(2):182–90.

20 Choi J, Johantgen M. The importance of supervision in retention of CNAs. Res Nurs Health 2012;35(2):187–99.

21 Loomer L, Grabowski DC, Yu H, et al. Association between nursing home staff turnover and infection control citation. Health Services Research 2021;57(2):322–32.

22 Molero-Jurado MDM, Pérez-Fuentes MDC, Gázquez-Linares, JJG, et al. Burnout risk and protection factors in certified nursing aides. Int J Environ Res Public Health 2018;15(6):1116.

23 Hagerty D, Buelow JR. Certified nursing assistants’ perceptions and generational differences. Am J Health Sci 2017;8(1):1–6.

24 Saiki M, Takemura Y, Kunie K. Nursing assistants’ desired roles, perceptions of nurses’ expectations and effect on team participation: a cross-sectional study. J Nurs Manag 2021;29(5):1046–53.

25 Cao J, Zhang L, Zhu J, et al. [The current situation of nursing staff grading at home and abroad, and assumption of nursing staff grading in China]. [Nursing Journal of Chinese People’s Liberation Army] 2006;(6):44–6.

26 Wang L, Guo H, Lei Y, et al. [Personnel staffing in long term-care facilities: a review]. [Chinese Journal of Nursing] 2014:49(8):981–5.

27 The Joanna Briggs Institute. The Joanna Briggs Institute reviewers’ manual 2015: methodology for JBI scoping reviews. Available: https://nursing.lsuhsc.edu/JBI/docs/ReviewersManuals/Scoping-.pdf [Accessed 3 Nov 2021].

28 Peters M, Godfrey C, Khalil H, et al. Guidance for conducting systematic scoping reviews. Int J Evid Based Healthc 2015;13(3):141–6.

29 Tricco AC, Lillie E, Zarin W, et al. PRISMA Extension for Scoping Reviews (PRISMA-ScR): Checklist and Explanation. Ann Intern Med 2018;169(7):467–73.

30 Zeng B, Du J. Administration models of education, training and appraisal of nursing assistants in inpatient care: a protocol for a scoping review. Available: https://doi.org/10.13140/RG.2.2.29106.12483/1 [Accessed 2 Nov 2021].

31 Moher D, Shamseer L, Clarke M, et al. Preferred reporting items for systematic review and meta-analysis protocols (PRISMA-P) 2015 statement. Syst Rev 2015;4(1):1.

32 Shamseer L, Moher D, Clarke M, et al. Preferred reporting items for systematic review and meta-analysis protocols (PRISMA-P) 2015: elaboration and explanation. Br Med J 2015;349:g7647.

33 Koster J. PubMed PubReMiner. Available: https://hgserver2.amc.nl/cgi-bin/miner/miner2.cgi [Accessed 31 Oct 2021].

34 Hong Q, Pluye P, Fàbregues S, et al. Mixed Methods Appraisal Tool (MMAT) version 2018. Available: http://mixedmethodsappraisaltoolpublic.pbworks.com/ [Accessed 7 Nov 2021].

35 Pluye P, Hong QN. Combining the power of stories and the power of numbers: mixed methods research and mixed studies reviews. Annu Rev Public Health 2014;35(1):29–45.

36 Shea BJ, Reeves BC, Wells G, et al. AMSTAR 2: a critical appraisal tool for systematic reviews that include randomised or non-randomised studies of healthcare interventions, or both. Br Med J 2017;358:j4008.

37 Sandelowski M, Barroso J, Voils CI. Using qualitative metasummary to synthesize qualitative and quantitative descriptive findings. Res Nurs Health 2007;30(1):99–111.

38 Appleby BE. Implementing guideline-checklists: Evaluating health care providers intentional behaviour using an extended model of the theory of planned behaviour. J Eval Clin Pract 2019;25(4):664–75.

39 Campbell AR, Kennerly S, Swanson M, et al. Manager’s influence on the registered nurse and nursing assistant relational quality and patient safety culture. J Nurs Manag 2021;29(8):2423–32.

40 Campbell AR, Layne D, Scott E, et al. Interventions to promote teamwork, delegation and communication among registered nurses and nursing assistants: an integrative review. J Nurs Manag 2020;28(7):1465–72.

41 Dutton S, Kozachik SL. Evaluating the outcomes of a web-based stress management program for nurses and nursing assistants. Worldviews Evid Based Nurs 2020;17(1):32–8.

42 Dykes PC, Carroll D, McColgan K, et al. Scales for assessing self-efficacy of nurses and assistants for preventing falls. J Adv Nurs 2011;67(2):438–49.

43 Feng R, Li J, Shi N, et al. [The comparison research of the working conditions of nursing assistants under directly and indirectly employed management models]. [Medicine and Society] 2013;26(10):39–41.

44 Friesen L, Andersen E. Outcomes of collaborative and interdisciplinary palliative education for health care assistants: a qualitative metasummary. J Nurs Manag 2019;27(3):461–81.

45 Gao J, Zhao H. [Dual management based on evidence-based nursing care in organizing nursing attending workers]. [Nursing of Integrated Traditional Chinese and Western Medicine] 2017;3(11):169–72.

46 Geoffrion S, Hills DJ, Ross HM, et al. Education and training for preventing and minimizing workplace aggression directed toward healthcare workers. Cochrane Database Syst Rev 2020;9(9):CD011860.

47 Haigh SM, Garside J. Effects of the Care Certificate on healthcare assistants’ ability to identify and manage deteriorating patients. Nurs Manage 2019;26(3):e1798.

48 Haraldsson P, Areskoug-Josefsson K, Rolander B, et al. Comparing the Structured Multidisciplinary work Evaluation Tool (SMET) questionnaire with technical measurements of physical workload in certified nursing assistants in a medical ward setting. Appl Ergon 2021;96:103493.

49 Kennerly SM, Yap TL, Hemmings A, et al. Development and psychometric testing of the nursing culture assessment tool. Clin Nurs Res 2012;21(4):467–85.

50 Lee C, Knight SW, Smith SL, et al. Safe patient handling and mobility: development and implementation of a large-scale education program. Crit Care Nurs Q 2018;41(3):253–63.

51 Liu J, Lin F, Feng Y, et al. [Management and practice of attending workers in hospital]. [Nursing of Integrated Traditional Chinese and Western Medicine] 2017;3(4):156–8.

52 Ma Y, Yin Y, Zhang Y. [FMEA method in the management of nursing workers in hospital]. [China Rural Health] 2019;11(15):20–1.

53 McKenzie SM, Turkhud K. Simulation-based education in support of HCA development. British Journal of Healthcare Assistants 2013;7(8):392-7. 54

54 Monteiro MS, Alexandre NMC, Milani D, et al. Work capacity evaluation among nursing aides. Revista da Escola de Enfermagem 2011;45(5):1177–82.

55 Nie Y, Dou L, Wang Y. [Application of group collaborative training model in field training for nursing assistants]. [Chinese Nursing Management] 2017;17(6), 808–10.

56 Nørgaard B, Ammentorp J, Kyvik KO, et al. Communication skills training increases self-efficacy of health care professionals. J Contin Educ Health Prof 2012;32(2):90–7.

57 Pfeifer P, Vandenhouten C, Purvis S, et al. The impact of education on certified nursing assistants’ identification of strategies to manage behaviors associated with dementia. J Nurses Prof Dev 2018;34(1):26–30.

58 Prestia A, Dyess S. Maximizing caring relationships between nursing assistants and patients: Care Partners. J Nurs Adm 2012;42(3):144–7.

59 Qiu Y, Wang J, Wu H, et al. [Practice and effects of construction of nursing assistant classification service standards based on Activities of Daily Living Scale]. [Chinese Journal of Modern Nursing] 2020;26(21):2858–62.

60 Ritchie R, Wood S, Martin FC, et al. Impact of an educational training program on restorative care practice of nursing assistants working with hospitalized older patients. J Clin Outcomes Manag 2017;24(9):425–32.

61 Small A, Atleno OL, Joseph T. Continuing education for patient care technicians: a unit-based, RN-led initiative. Am J Nurs 2012;112(8):51–5.

62 Swann RA. The Effect of Training and Recognition on Nursing Assistant Retention in Acute Care Settings [master’s dissertation]. Cleveland: Gardner-Webb University; 2018. 46 p.

63 Tom C. Improving Patient Safety through Patient Safety Aide (Sitter) Competency Education [doctor’s dissertation]. Cleveland: Gardner-Webb University; 2016. 80 p.

64 Twigg DE, Myers H, Duffield C, et al. The impact of adding assistants in nursing to acute care hospital ward nurse staffing on adverse patient outcomes: an analysis of administrative health data. Int J Nurs Stud 2016;63:189–200.

65 Wagner EA. Improving patient care outcomes through better delegation-communication between nurses and assistive personnel. J Nurs Care Qual 2018;33(2):187–93.

66 Ward S, Stewart D, Ford D, et al. Educating certified nursing assistants educational offerings on the run and more. J Nurses Prof Dev 2014;30(6):296–302.

67 Wilson CM, Baumont MM, Alberstadt KM, et al. The effectiveness of a patient handling education program for nursing assistants as taught by physical therapy and nursing educators. J Acute Care Phys Ther 2011;2(1):12–38.

68 Wu L, Pu M, Chen Y. [Application of hospital management model to the management of nurse attendants]. [Journal of Qilu Nursing] 2015;21(12):21–3.

69 Yu Q. [Application of Quality Control Circle to hand hygiene of nursing assistants in psychiatric geriatric wards]. [Journal of Traditional Chinese Medicine Management] 2015;23(8):58–60.

70 Zhao W, Wang Y, Han J, et al. [Effects of stratified training of nursing assistants based on patient demands]. [Chinese Journal of Modern Nursing] 2020;26(32):4533–6.

71 Zhi M, He Z, Ji J, et al. Patient satisfaction with non-clinical nursing care provided by the nursing assistant under different management models in Chinese public tertiary hospital. Appl Nurs Res 2021;151431.

72 Zhu D, Jiang Y, Chen Y, et al. [Practice of nursing assistant self-protection training program during the epidemic period of COVID-19]. [Shanghai Nursing] 2021;21(9):61–3.

73 Zhu F, Qu X, Hu C. [Effectiveness of the inform sheet in the management of nursing assistants]. [China Health Industry] 2019;16(32):125–6.

74 Rockwell SK, Kohn H. Post-then-pre evaluation: Measuring behavior change more accurately. J Ext 1989;27:19–21.

75 Agency for Healthcare Research and Quality. TeamSTEPPS 2.0. Available: https://www.ahrq.gov/teamstepps/instructor/index.html [Accessed 26 Nov 2021].

76 Graen GB, Uhl-Bien M. Relationship-based approach to leadership: development of leader-member exchange (LMX) theory of leadership over 25 years: applying a multi-level multi-domain perspective. Leadersh Q 1995;6(2):219–47.

77 Haraldsson P, Jonker D, Strengbom E, et al. Structured Multidisciplinary work Evaluation Tool: development and validation of a multidisciplinary work questionnaire. Work 2016;55:883–91.

78 Ilmarinen J. The work ability index (WAI). Occup Med 2007;57(2):160.

79 Chen L, Cheng Y, Meng M, et al. [Analysis of the nursing assistant management models in China]. [Modern Nurse] 2020;27(19):21–2.

80 Li G, Ying Y. [Current situations of attendant management]. [Modern Hospitals] 2021;21(5):739–41.

81 Song J. Study on the situation, problem and strategies about nursing assistants in health institutions of Z province [master’s dissertation]. Hangzhou: Zhejiang University; 2016, 60 p.

82 Wang Y, Jia T, Yuan H. [International comparison of the nursing assistants industry]. [Chinese Hospitals] 2016;20(11):76–8.

83 Asgari-Dastjerdi H, Khorasani E, Yarmohammadian, MH, et al. Evaluating the application of failure mode and effects analysis technique in hospital wards: a systematic review. J Inj Violence Res 2017, 9(1), 51–60.

84 Liu HC, Zhang LJ, Ping YJ, et al. Failure mode and effects analysis for proactive healthcare risk evaluation: a systematic literature review. J Eval Clin Pract 2020;26(4):1320–37.

85 Gilly BA, Touran A, Asai T. Quality control circles in construction. J Constr Eng Manag 1987;113(3):427–39.

86 Rohrbasser A, Harris J, Mickan S, et al. Quality circles for quality improvement in primary health care: Their origins, spread, effectiveness and lacunae-A scoping review. PLoS One 2018;13(12):e0202616.

87 The Staff of the Benjamin Rose Hospital. Multidisciplinary studies of illness in aged persons: II. A new classification of functional status in activities of daily living. J Chronic Dis 1959;9(1):55–62.

88 Huckabay, L. M. (1991). The role of conceptual frameworks in nursing practice, administration, education, and research. Nur Adm Q 1991;15(3):17–28.

89 Cook DA, Hatala R, Brydges R, et al. Technology-enhanced simulation for health professions education: a systematic review and meta-analysis. JAMA 2011;306(9):978–88.

90 Mulyadi M, Tonapa SI, Rompas, SSJ, et al. Effects of simulation technology-based learning on nursing students’ learning outcomes: a systematic review and meta-analysis of experimental studies. Nurse Educ Today 2021;107:105127.

91 Lee J, Lee H, Kim S, et al. Debriefing methods and learning outcomes in simulation nursing education: a systematic review and meta-analysis. Nurse Educ Today 2020;87:104345.

92 Al-Balas M, Al-Balas HI, Jaber HM, et al. Distance learning in clinical medical education amid COVID-19 pandemic in Jordan: current situation, challenges, and perspectives. BMC Med Educ 2020;20(1):341.

93 Rouleau G, Gagnon MP, Côté J, et al. Effects of e-learning in a continuing education context on nursing care: systematic review of systematic qualitative, quantitative, and mixed-studies reviews. J Med Int Res 2019;21(10):e15118.

94 Voutilainen A, Saaranen T, Sormunen M. Conventional vs. e-learning in nursing education: a systematic review and meta-analysis. Nurse Educ Today 2017;50:97–103.

95 Bowers B, Becker M. Nurse’s aides in nursing homes: the relationship between organization and quality. Gerontologist 1992;32(3):360–6.

96 D’Arcy LP, Sasai Y, Stearns SC. Do assistive devices, training, and workload affect injury incidence? Prevention efforts by nursing homes and back injuries among nursing assistants. J Adv Nurs 2012;68(4):836–45.

97 Epstein NE. Multidisciplinary in-hospital teams improve patient outcomes: a review. Surg Neurol Int 2014;5(8):S295–303.

98 Squires M, Tourangeau A, Spence LHK, et al. The link between leadership and safety outcomes in hospitals. J Nurs Manag 2010;18(8):914–25.

