## Supplementary material for "Administration approaches of nursing assistants in hospitals: a scoping review": online supplemental file 1

### **Supplemental File 1** Full search strategies

**S1.1** PubMed, search completed on November 7, 2021; updated on March 18, 2022

| # | Query | Records (Nov 7, 2021) | Records (Mar 18, 2022) |
|---|---|---|---|
| 1 | "nursing assistants"[mh] | 4,714 | 4,742 |
| 2 | "nursing assistant*"[tiab] OR "healthcare assistant*"[tiab] OR "health care assistant*"[tiab] OR "nurse assistant*"[tiab] OR "nurse aide*"[tiab] OR "nursing aide*"[tiab] OR "nursing auxiliar*"[ti] OR "care aide*"[ti] OR "paramedic*"[ti] OR "carer"[ti] OR ("nurs*"[ti] AND "aides"[ti]) | 7,555 | 7,746 |
| 3 | #1 OR #2 | 10,807 | 11,003 |
| 4 | "educat*"[ti] OR "training"[ti] OR "evaluat*"[ti] OR "assess*"[ti] OR "exam*"[ti] OR "apprais*"[tiab] OR "organisation"[ti] OR "health care quality"[tiab] OR "healthcare quality"[ti] OR "framework"[tiab] OR "administr*"[ti] OR ("management"[ti] AND "model"[ti]) OR "clinical governance"[mh] OR "nursing administration research"[mh] | 1,885,997 | 1,934,139 |
| 5 | "standards"[sh] OR "education"[sh] OR "education"[mh] OR "organization and administration"[sh] OR "organization and administration"[mh] | 3,507,001 | 3,564,605 |
| 6 | #4 AND #5 | 502,640 | 512,125 |
| 7 | "clinical trial protocol"[pt] OR "review"[pt] OR "case reports"[pt] | 4,998,699 | 5,076,927 |
| 8 | "protocol"[ti] OR "student*"[ti] OR "home*"[ti] OR "long-term*"[ti] OR "long term*"[ti] OR "outpatient"[ti] | 555,696 | 571,296 |
| 9 | #7 OR #8 | 5,492,143 | 5,594,066 |
| 10 | "2011/01/01"[pdat]:"3000/12/31"[pdat] | 12,511,650 | 647,617* |
| 11 | #3 AND #6 AND #10 NOT #9 | 402 | 4 (4 added) |

*: #10 searched on Mar 18, 2022 using expression “"2021/11/08"[pdat]:"3000/12/31"[pdat]”

**S1.2** Scopus, search completed on November 7, 2021; updated on March 19, 2022

| # | Query | Records (Nov 7, 2021) | Records (Mar 19, 2022) |
|---|---|---|---|
| 1 | TITLE-ABS-KEY ("nursing assistant*" OR "healthcare assistant*" OR "health care assistant*" OR "nurse assistant*" OR "nurse aide*" OR "nursing aide*") | 8,337 | 8,532 |
| 2 | TITLE ("nursing auxiliar*" OR "care aide*" OR "paramedic*") OR TITLE ("nurs*" PRE/0 "aide*") | 4,353 | 4,449 |
| 3 | #1 OR #2 | 11,991 | 12,273 |
| 4 | TITLE ("educat*" OR "training" OR "evaluat*" OR "assess*" OR "exam*" OR "apprais*" OR "organisation" OR "health care quality" OR "healthcare quality" OR "framework" OR "administr*" OR ("management" W/5 "model") OR "clinical governance" OR "nursing administration research") | 4,050,243 | 4,157,068 |
| 5 | ABS ("clinical governance" OR "nursing administration" OR "nursing management") OR KEY ("framework" OR "apprais*") | 350,701 | 358,440 |
| 6 | #4 OR #5 | 4,276,751 | 4,391,873 |
| 7 | DOCTYPE (ar OR ab OR bk OR ch OR sh) | 63,429,512 | 64,546,137 |
| 8 | SUBJAREA (medi OR nurs OR heal) OR SUBJAREA (busi OR deci OR econ OR psyc OR soci) | 36,674,404 | 37,333,199 |
| 9 | #7 AND #8 | 28,273,499 | 28,788,473 |
| 10 | INDEX (medline) | 28,297,255 | 28,772,100 |
| 11 | TITLE ("protocol" OR "student*" OR "home*" OR "long-term*" OR "long term*" OR "outpatient" OR "school") | 1,386,755 | 1,425,899 |
| 12 | #10 OR #11 | 29,115,435 | 29,613,965 |
| 13 | #9 AND NOT #12 | 11,788,807 | 12,068,017 |
| 14 | PUBYEAR > 2010 | 33,766,020 | 4,623,945* |
| 15 | LANGUAGE (English OR Chinese) | 75,230,770 | 76,699,943 |
| 16 | #3 AND #6 AND #13 AND #14 AND #15 | 160 | 43 (17 added)† |

*: #14 searched on Mar 19, 2022 using expression “PUBYEAR AFT 2020”

†: from the 43 articles searched, 17 were published after the initial searching date (November 7, 2021).

**S1.3** CINAHL, search completed on November 8, 2021; updated on March 19, 2022

| # | Query | Records (Nov 8, 2021) | Records (Mar 19, 2022) |
|---|---|---|---|
| S1 | MH ("nursing assistants") | 8,587 | 8,662 |
| S2 | TI ("nursing assistant*" OR "healthcare assistant*" OR "health care assistant*" OR "nurse assistant*" OR "nurse aide*" OR "nursing aide*" OR "nursing auxiliar*" OR "care aide*" OR "paramedic*" OR "carer") OR AB ("nursing assistant*" OR "healthcare assistant*" OR "health care assistant*" OR "nurse assistant*" OR "nurse aide*" OR "nursing aide*") | 8,820 | 9,011 |
| S3 | S1 OR S2 | 15,048 | 15,275 |
| S4 | TI ("educat*" OR "training" OR "evaluat*" OR "assess*" OR "exam*" OR "organisation" OR "healthcare quality" OR "administr*" OR "apprais*" OR "health care quality" OR "framework") OR TI ("management" AND "model") OR AB ("apprais*" OR "health care quality" OR "framework") | 628,913 | 646,769 |
| S5 | MH ("clinical governance" OR "nursing administration research") | 2,570 | 2,615 |
| S6 | S4 OR S5 | 630,598 | 648,876 |
| S7 | MW ("quality" OR "education" OR "administration" OR "organization") OR TI ("quality" OR "education" OR "administration" OR "organization") OR MH ("management") | 1,413,116 | 1,437,840 |
| S8 | S6 AND S7 | 256,302 | 262,174 |
| S9 | PT ("review" or "protocol") | 342,946 | 351,021 |
| S10 | TI ("protocol" OR "student*" OR "home*" OR "long-term*" OR "long term*" OR "outpatient") | 265,762 | 273,860 |
| S11 | S9 OR S10 | 599,774 | 615,058 |
| S12 | S3 AND S8 NOT S11 | 1,072 | 1,083 |
| | Limiters: Published Date: 20110101-20250131; English Language; Scholarly (Peer Reviewed) Journals; Research Article; Exclude MEDLINE records; Language: English;<br>OR<br>Limiters: Published Date: 20110101-20250131; English Language; Exclude MEDLINE records; Language: English; Source Types: Dissertations | 216 | 8 (8 added) * |

*: Limiters for updated searching were: Published Date: 20211101-20251231; English Language; Scholarly (Peer Reviewed) Journals; Research Article; Exclude MEDLINE records; Language: English

**S1.4** CNKI, WanFang Med, and SinoMed, search completed on November 7, 2021; updated on March 19, 2022

| Database | Query | Records (Nov 7, 2021) | Records (Mar 19, 2022) |
|---|---|---|---|
| CNKI | (TI=('护理员'+'护工'+'助理护士') OR KY=('护理员'+'护工'+'助理护士')) AND ((TI=('管理'+'培训'+'评价'+'考核')) OR (KY=('管理'+'培训'+'评价'+'考核'))) NOT TI=('养老') AND TKA=('模式'+'问卷'+'量表'+'研究'+'质性'+'调查'+'访谈') | 137 | 143 |
| | Limit to: 资源范围:学术期刊,学位论文,会议；时间范围：发表时间:2011-01-01 到；更新时间:不限。文献分类:医药卫生科技,社会科学Ⅱ辑,经济与管理科学 | 117 | 1 (1 added) * |
| WanFang | ((主题="护理员" OR "护工" OR "助理护士") AND 题名或关键词=("管理" OR "培训" OR "评价" OR "考核")) AND 题 | 323 | 333 |

| | | | |
|---|---|---|---|
| Med | 名或关键词=("模式" OR "问卷" OR "量表" OR "质性" OR "调查" OR "访谈" OR "试验" OR "实验") | | |
| | Limit to: 年份=2011-2021 资源类型=(中文期刊 OR 学位论文 OR 会议论文) | 256 | 20 (1 added) † |
| SinoMed | (("助理护士"[标题] OR "护理员"[标题] OR "护工"[标题] OR "助理护士"[不加权:不扩展]) AND ("管理"[标题] OR "培训"[标题] OR "评价"[标题] OR "考核"[标题] OR "护理管理研究"[不加权:不扩展] OR "管理模式"[摘要] OR "管理体系"[摘要] OR "胜任力"[摘要] OR "职业倦怠"[摘要]) AND ("模式"[标题] OR "问卷"[标题] OR "量表"[标题] OR "研究"[标题] OR "质性"[标题] OR "调查"[标题] OR "访谈"[标题] OR "质性"[摘要] OR "问卷"[摘要] OR "对照"[摘要] OR "循证"[摘要])) AND ("2021"[时间] OR "2020"[时间] OR "2019"[时间] OR "2018"[时间] OR "2017"[时间] OR "2016"[时间] OR "2015"[时间] OR "2014"[时间] OR "2013"[时间] OR "2012"[时间] OR "2011"[时间]) NOT "养老"[标题] | 160 | 11 (3 added) ‡ |

*: Limiters for updated searching were: publication time later than 2021-11-07.

†: Limiters for updated searching were: publication year = 2021-2022; 1 of the 20 records was published after the initial searching date (Nov 7, 2021).

‡: Publication year for updated searching was adjust to “2021 OR 2022”; 3 of the 11 records was published after the initial searching date (Nov 7, 2021).

**S1.5** NICE, AHRQ, and CADTH, search completed on November 8, 2021; updated on March 19, 2022

| Database | Query | Records (Nov 8, 2021) | Records (Mar 19, 2022) |
|---|---|---|---|
| NICE: NHS Evidence | ("health care assistant*" OR "nursing assistant*" OR "healthcare assistant*" OR "nursing aide*") AND ("educat*" OR "training" OR "evaluat*" OR "assess*" OR "exam*" OR "organisation" OR "healthcare quality" OR "administr*") | 365 | - |
| | Filter: From 2011/01/01 to 2021/11/08, Evidence Type: policy and strategy, guidance, prescribing and technical information, systematic reviews, quality indicators | 119 | 8 (8 added) * |
| AHRQ | Limit: Searched titles only | | |
| | "nursing assistant" (2 records), "nursing aide" (0 record), "care aide" (1 record), "healthcare assistant" (0 record) | 3 | 0 (0 added) |
| CADTH | "nursing assistant"：No records available. | 9, but 0 available | 0 (0 added) |

*: Filter for additional searching was adjust to “From 2021/11/08 to 2022/03/19, All Evidence type”.

**S1.6** ProQuest, search completed on November 8, 2021; updated on March 19, 2022

| # | Query | Records (Nov 8, 2021) | Records* (Mar 19, 2022) |
|---|---|---|---|
| S1 | MESH("nursing assistants") | 322 | - |
| S2 | TI,AB("nursing assistant*" OR "healthcare assistant*" OR "health care assistant*" OR "nurse assistant*" OR "nurse aide*" OR "nursing aide*") OR TI("nursing auxiliar*" OR "care aide*" OR "paramedic*" OR "carer") OR TI("nurs*" PRE/0 "aide*") | 6,707 | - |
| S3 | 1 or 2 | 6,891 | - |
| S4 | TI("educat*" OR "training" OR "evaluat*" OR "assess*" OR "exam*" OR "organisation" OR "healthcare quality" OR "administr*") OR TI("management" W/3 "model") OR TI,AB("apprais*" OR "health care quality" OR "framework") | 2,463,235 | - |
| S5 | MESH("clinical governance" OR "nursing administration research") | 199 | - |
| S6 | 4 or 5 | 2,463,413 | - |
| S7 | SU,TI("standards" OR "education" OR "organization and administration") OR MESH("education" OR "organization and administration") | 3,060,345 | - |
| S8 | 6 and 7 | 1,034,055 | - |
| S9 | YR(2011-3000) | 11,947,480 | † |
| S10 | 3 AND 8 AND 9 | 143 | 13 (13 added) |

the following databases were searched by ProQuest on November 8, 2021: Health & Medical Collection, Social Science Collection, and ProQuest Dissertations & Thesis. ERIC was retrieved by ProQuest.

*: Due to the subscription and access change of the ProQuest Databases, searching on March 19, 2022 used different databases (56 databases).

†: Filter “publication date 2021-11-08 - 2022” was used for the update searching.

**S1.7** JBI EBP, Cochrane DSR, Embase, Emcare and PsycInfo via Ovid, search completed on November 7, 2021; updated on March 19, 2022

| # | Query | Records (Nov 7, 2021) | Records (Mar 19, 2022) |
|---|---|---|---|
| 1 | "nursing assistants".mh. | 39 | 42 |
| 2 | ("nursing assistant*" OR "healthcare assistant*" OR "health care assistant*" OR "nurse assistant*" OR "nurse aide*" OR "nursing aide*").ti,ab. OR ("nursing auxiliar*" OR "care aide*" OR "paramedic*" OR "carer").ti,kw. OR ("nurs*" AND "aides").ti. | 18,502 | 19,051 |
| 3 | 1 or 2 | 18,516 | 19,066 |
| 4 | ("educat*" OR "training" OR "evaluat*" OR "assess*" OR "exam*" OR "organisation" OR "healthcare quality" OR "administr*").ti,kw. OR ("management" AND "model").ti. OR ("apprais*" OR "health care quality" OR "framework").ti,ab. | 3,553,150 | 3,643,169 |
| 5 | ("clinical governance" OR "nursing administration research").mh. | 395 | 395 |
| 6 | 4 or 5 | 3,553,468 | 3,643,487 |
| 7 | ("standards" OR "education" OR "organization and administration").sh. OR ("education" OR "organization and administration").mh. | 705,599 | 716,162 |
| 8 | 6 and 7 | 215,019 | 219,032 |
| 9 | ("conference review" OR "erratum" OR "letter" OR "note" OR "protocol" OR "review" OR "systematic review protocols").pt. | 6,620,263 | 6,756,969 |
| 10 | ("protocol" OR "student*" OR "home*" OR "long-term*" OR "long term*" OR "outpatient" OR "school").ti,kw. | 1,346,568 | 1,381,671 |
| 11 | 9 or 10 | 7,849,293 | 8,017,286 |
| 12 | (3 and 8) not 11 | 668 | 684 |
| 13 | Filter: Specific Year Range, from 2011 to 5000. | 326 | 35 (16 added)* |

*: Filter for updated searching was adjust to “Specific Year Range, from 2021 to 5000”; 16 of the 35 records was published after the initial searching date (Nov 7, 2021).
