## Supplementary material for "Administration approaches of nursing assistants in hospitals: a scoping review": online supplemental file 2

**Supplemental File 2** Study Selection Checklist for Screening

S2.1 For title screening:

| If contains | of the items: | the study should be |
|---|---|---|
| any | (a) nursing assistant as a subject. | included |
| | (b) nursing-related personnel as a subject (nursing assistants were included). | |
| | (c) models, frameworks and theories. | |
| | (d) proper nouns of theories, models, approaches or scales. | |
| any | (a) patient as a subject. | excluded |
| | (b) knowledge on specific diseases or symptoms, and not generally applicable. | |
| | (c) review (no evidence-based). | |
| | (d) nursing homes, nursing house, community care or long-term care facilities. | |
| | ***NOT*** any related description on nursing workers and administration, ***and*** describes a completely unrelated topic | excluded |
| | ***neither*** any related description on nursing workers and administration, ***nor*** any unrelated description | included for further screening |

S2.2 For abstract screening:

| If contains | of the items: | the study should be |
|---|---|---|
| all | (a) Nursing assistants were included in the study, and were independently displayed | included |
| | (b) Any reasonable methodologies. | |
| | (c) Clear and detailed interventions or exposures that conformed to the topics. | |
| | (d) Any outcomes that proved significance or negative results. | |
| any | (a) Paramedics, healthcare staffs, or other healthcare workers as an entire subject. | excluded |
| | (b) No details of methodologies, interventions or exposures. | |
| | (c) Interventions, exposures or outcomes that were only fit for a specific department, disease or field. | |
