## Supplementary material for "Administration approaches of nursing assistants in hospitals: a scoping review": online supplemental file 3

**Supplemental File 3** A Data Charting (Data Extraction) Tool

***Directions*** This is a data charting tool for the scoping review, organized as fourths: study characteristics, participants & methodologies, outcomes, and a data charting checklist for laws.

1. Full text of the publications should be read carefully before data charting, and extracting data from abstract were not allowed.
2. Identify study types carefully, for wrong study types may lead to wrong selection of tools.
3. If failing to determine what to chart or leave, a third-reviewer discussion will resolve the confusion ultimately. Please mark the study and wait for discussion.

***Part 1: Study Characteristics***

| | | | | |
|---|---|---|---|---|
| Title | | | | |
| Type | ○research paper ○evidence synthesis ○dissertation ○others ________ | | | |
| Author | | Origin | | Pub Year |
| Area | ○medicine ○nursing ○pub health ○social science ○jurisprudence | | | |
| Study Type | | ○qualitative ○quantitative | | ○prospective ○retrospective |
| | | ○descriptive ○interventional | | evidence-based: ○yes ○no |
| | | ○tool assessment | | ○others |
| | | specific type: ________________________________ | | |
| Practice Settings | | | | |
| Topic | ○administration ○education ○training ○appraisal ○selection ○others | | | |
| | specific topic: ______________________________________________ | | | |

***Part 2: Participants & Methodologies***

| | | | | | | |
|---|---|---|---|---|---|---|
| Participants | ○only nursing assistants ○mixed, nursing ○mixed, healthcare | | | | | |
| | Definition | | | | | |
| | Characteristics | Number | | Age | | Gender R |
| | | Educ BKG | | Ethnicity | | |
| | Demographic | | | | | |
| | Response R. | | | | | |
| Methodology | Theories | | Statistics | | | |
| | Interventions / Exposures | | Comparators | | | |
| | Duration | | Measure / Appraisal | | | |
| | Others: ____________________________________________________ | | | | | |

***Part 3: Outcomes***

| | | |
|---|---|---|
| First Outcome | Topic | ○administration ○education ○training ○appraisal ○selection |
| | Type | ○theory ○approach ○model ○effectiveness ○tool ○others |
| | Outcome | |
| | Statistic Data | |
| Second Outcome | Topic | ○administration ○education ○training ○appraisal ○selection |
| | Type | ○theory ○approach ○model ○effectiveness ○tool ○others |
| | Outcome | |
| | Statistic Data | |
| Key Findings | | |
| Others | | |

***Part 4: Others (if applicable)***

| |
|---|
| Limitations |
| Fundings |
| Conflicts of Interest |
