## Supplementary material for "Administration approaches of nursing assistants in hospitals: a scoping review": online supplemental file 4

**Supplemental File 4** Reason for exclusion (*n* = 112)

| No. | Publication | Excluded reason |
|---|---|---|
| 1 | Applying Kane's validity framework to a simulation based assessment of clinical competence (Tavares et al., 2018) | participants were mixed and NAs were not displayed and evaluated separately. |
| 2 | Assessing nursing assistants' competency in palliative care: An evaluation tool (Karacsony et al., 2018) | setting: aged care facilities. |
| 3 | Assessment of patient safety culture among paramedical personnel at general and district hospitals, Fayoum Governorate, Egypt (El-Sherbiny et al., 2020) | participant: paramedical staff, not including NAs. |
| 4 | Associations between state regulations, training length, perceived quality and job satisfaction among certified nursing assistants: cross-sectional secondary data analysis (Han et al., 2014) | setting: data from NNHS and NNAS were on nursing homes. |
| 5 | Can an interprofessional education tool improve healthcare professional confidence, knowledge and quality of inpatient diabetes care: a pilot study? (Herring et al., 2013) | specific area: conclusions were only applied to diabetes care. |
| 6 | Carer assessment: continuing tensions and dilemmas for social care practice (Seddon & Robinson, 2015) | participant: carers, not including NAs. |
| 7 | Certified Nursing Assistants Want to Use External Memory Aids for Residents With Dementia: Survey Results Within an Implementation Science Framework (Douglas & Affoo, 2019) | main focus and specific area: conclusions were only applied to dementia care, and main focus was not on NA administration |
| 8 | The challenges of training, support and assessment of healthcare support workers: A qualitative study of experiences in three English acute hospitals (Sarre et al., 2018) | main focus: the study was descriptive and did not focus on NA administration approaches. |
| 9 | Choosing Educational Resources to Build Interprofessional, Palliative Care Competency: A Replicable Review Methodology (Yue & Tayler, 2020) | type: a literature review |
| 10 | Communication skills training in end-of-life care (Morris, 2011) | methodology: methodology was not described. |
| 11 | *Communication with patients and colleagues: An intervention study on the impact of a communication skills training course on health care professionals' ability to communicate with patients and colleagues (Nørgaard, 2011) | full text not found. |
| 12 | *"Compulsory training would help every HCA spot moisture lesions" (Elson, 2011) | full text not found. |
| 13 | ("Council's report to members on Congress 2013,") | type: a report from Royal College of Nursing. |
| 14 | Current status of clinical education in paramedic programs: A descriptive research project (Grubbs, 2014) | publication year was 1997, but retrieved from the database. |
| 15 | Educating Certified Nursing Assistants to Communicate Skin Changes to Reduce Pressure Injuries (Rummel et al., 2021) | setting: long-term care setting, not a hospital. |
| 16 | Education for healthcare assistants working in acute NHS hospitals (Webb, 2011) | type: a literature review. |
| 17 | Education for healthcare clinical support workers (Lewis & Kelly, 2015) | type: a literature review. |
| 18 | An Educational Intervention to Improve Staff Collaboration and Enhance Knowledge of Fall Risk Factors and Prevention Guidelines (DiGerolamo & Chen-Lim, 2021) | participants (RNs and NAs) were mixed and NAs were not separately discussed. |
| 19 | The effect of an e-learning course on nursing staff's knowledge of delirium: A before-and-after study (Van De Steeg et al., 2015) | specific area: outcomes were only fit for delirium care |
| 20 | The Effectiveness and Need for Facility Based Nurse Aide Training Competency Evaluation Programs (Mileski et al., 2016) | setting: data from NNAS were on nursing homes. |
| 21 | Effectiveness of an Educational Intervention With High-Fidelity Clinical Simulation to Improve Attitudes Toward Teamwork Among Health Professionals (Muñoz de Morales-Romero et al., 2021) | participants (all health care professionals) were mixed and NAs were not separately discussed. |
| 22 | Effects of an advanced nursing assistant education program on job satisfaction, turnover rate, assistant education program on and clinical outcomes (Brown et al., 2013) | setting: long-term care. |
| 23 | The Effects of Education About Urinary Incontinence on Nurses' and Nursing Assistants' Knowledge, Attitudes, Continence Care Practices, and Patient Outcomes: A Systematic Review (Ostaszkiewicz et al., 2020) | specific area: this systematic review was on rehabilitation care. |
| 24 | The effects of executive involvement, goal setting, targeted education and caregiver recognition on hand hygiene performance (Bailey, 2013) | conference abstract. |
| 25 | Empowering CNAs With Training: Quality Outcomes for Nursing Facilities (Himes-Bissonnette, 2019) | setting: nursing home. |
| 26 | Evaluating the impact of pluridisciplinary training on proper glove use in hospital (Turco et al., 2014) | participants were mixed (nurses, nurse aides and housekeepers) and NAs were not evaluated separately. |
| 27 | Evaluating the success of the practical nursing students who are certified nurse aides (Stocking, 2012) | participant: NA students. |
| 28 | Examining Certified Nursing Assistants' Perceptions of Work-Related Identity (Gray & Lukyanova, 2017) | full text not found. |
| 29 | *The future education of nurses and healthcare assistants (Glasper, 2015) | methodology: the article did not show methodology. |
| 30 | General practitioners, primary care and support for carers in England: can training make a difference? (Jones et al., 2012) | participant: general practitioners. |
| 31 | A guide for HCAs on safe patient transfers (Lees, 2013) | methodology: the article did not show methodology. |
| 32 | How training could improve the way healthcare assistants relate to older people (Maxwell, 2018) | outcome: the article did not show the outcome. |
| 33 | Identifying a practice-based implementation framework for sustainable interventions for improving the evolving working environment: Hitting the Moving Target Framework (Højberg et al., 2018) | participant: the article was not for NAs. |
| 34 | Improving Retention Among Certified Nursing Assistants Through Compassion Fatigue Awareness and Self-Care Skills Education (Dreher et al., 2019) | setting: long-term care. |
| 35 | *("Improving the lives of carers," 2015) | full text not found. |
| 36 | Instructors for on-the-job training of advanced paramedics - definition of competencies and development of a quality management tool for a "High Responsibility Organization" (Flentje et al., 2019) | participant: trainees and practical instructors. |
| 37 | Measuring Carer Outcomes in an Economic Evaluation: A Content Comparison of the Adult Social Care Outcomes Toolkit for Carers, Carer Experience Scale, and Care-Related Quality of Life Using Exploratory Factor Analysis (Engel et al., 2020) | participant: carers, not including NAs. |
| 38 | *Minimum training standards for HCAs (Entwistle, 2013) | full text not found. |
| 39 | MULTI-DISCIPLINARY SIMULATION TRAINING ON DELIRIUM...British Geriatrics Society Autumn Meeting, November 25-27 2020 (Virtual) (Varma et al., 2021) | conference abstract. |
| 40 | Navigating the grounded theory terrain. Part 2 (Hunter et al., 2011) | main focus: about theory not on NA administration. |
| 41 | Needs Assessments to Determine Training Requirements (Lepicki & Boggs, 2014) | main focus: a need assessment project, not on NA administration approach. |
| 42 | Nurses' and nursing assistants' emotional skills: A major determinant of motivation for patient education (Lelorain et al., 2019) | participants were mixed (RNs and NAs) and NAs were not separately discussed. |
| 43 | ("Nursing Workforce Standards," 2021) | type: this is not a study so effectiveness cannot be identified. |
| 44 | On the Assessment of Paramedic Competence: A Narrative Review with Practice Implications (Tavares & Boet, 2016) | type: literature review. |
| 45 | A preliminary qualitative evaluation of the Virginia gold quality improvement program (Craver & Burkett, 2012) | setting: nursing home. |
| 46 | Psychometric testing of an instrument measuring nurse aides' patient safety attitudes (Perng & Yu, 2013) | setting: long-term care. |
| 47 | The relationship of positive work environments and workplace injury: evidence from the National Nursing Assistant Survey (McCaughey et al., 2014) | setting: data from NNAS were on nursing homes. |
| 48 | *Remit of training review revealed (Lintern, 2014) | full text not found |
| 49 | SIMULATED LEARNING FOR NURSING ASSISTANT EDUCATION IN ONTARIO, CANADA (Flynn et al., 2017) | setting: home care. |
| 50 | *Simulation in Nursing Education: Current Regulations and Practices (Hayden et al., 2014) | participant: RNs, practical and vocational nurses, not for NAs. |
| 51 | A study of the impact of three day training programme on knowledge regarding biomedical waste among paramedical staff of District hospital Etawah (UP) (Srivastava et al., 2013) | participants were mixed (paramedical staff) and NAs were not discussed separately. |
| 52 | Tailoring an information skills programme for Trainee Nursing Associates (Froste, 2020) | participant: trainee nursing associates, whose definition was not conformed to this review. |
| 53 | Testing carer skill training programs in Spanish carers of patients with eating disorders (Quiles Marcos et al., 2018) | participant: carers, and NAs were not included. |
| 54 | *Training and other important needs for nursing assistants (Robinson, 2011) | setting: long-term care. |
| 55 | *Training curriculum and simulator training for the whole surgical team: what do nurse assistants think? (Rosqvist et al., 2012) | type: a letter to the editor. |
| 56 | Training new HCAs to give compassionate care (Morgan et al., 2015) | methodology: the article did not include specific methodology. |
| 57 | *Training review will look into HCA role and path to becoming a nurse (Kleebauer, 2014) | type: news. |
| 58 | Undertaking special observation of patients with neurological conditions: evaluation of a training programme for HCAs (Flynn et al., 2016) | specific area: this article was only for patients with neurological diseases. |
| 59 | *Unqualified support staff are doing more observations with minimal training. Do standards exist for this work? (Lees, 2011) | full text not found. |
| 60 | Use of technologies in programs of permanent education in health: an experimental study (Neves Silva & Cordeiro, | outcome: the article did not perform outcomes. |

**Supplemental File 4** Reason for exclusion ($n$ = 112)

| No. | Publication | Excluded reason |
|---|---|---|
| | 2013) | |
| 61 | Utilizing Vroom's expectancy theory as a predictor of student academic success on the Illinois nurse assistant competency examination (Whittington, 2014) | main focus: this thesis was on the Illinois NA Training Competency Examination. |
| 62 | A Video-Based Intervention on and Evaluation of Nursing Aides' Therapeutic Communication and Residents' Agitation During Mealtime in a Dementia Care Unit (Levy-Storms et al., 2016) | setting: long-term care. |
| 63 | *A vital part of the team Erin Dean looks ahead at training for healthcare assistants (Dean, 2014) | type: this was not a research paper. |
| 64 | ("What Really Matters? A Vision for Nursing in Older People's Services," 2016) | participants were mixed (healthcare staff, service users, carers, and education providers) and NAs were not discussed separately. |
| 65 | The Work Organisation Assessment Questionnaire: validation for use with community nurses and paramedics (Karimi & Oakman, 2020) | participant: paramedics, not including NAs. |
| 66 | **Can Health-care Assistant Training improve the relational care of older people?:(CHAT) A development and feasibility study of a complex intervention (Arthur et al., 2017) | outcome: no outcomes on administration. |
| 67 | **The Cavendish Review (Cavendish, 2013) | type: most of this publication was not serious research. |
| 68 | **The changing boundaries of nursing: a qualitative study of the transition to a new nursing care delivery model (Rhéaume et al., 2015) | main focus: not on NAs. |
| 69 | **Empowering certified nurse's aides to improve quality of work life through a team communication program (Howe, 2014) | setting: long-term care. |
| 70 | **The first year: employment patterns and job perceptions of nursing assistants in a rural setting (MEYER et al., 2014) | setting: long-term care. |
| 71 | **Job Satisfaction of Nursing Assistants (Lerner et al., 2011) | main focus: this article was not on administration. |
| 72 | **Transformational leadership and workplace injury and absenteeism: Analysis of a national nursing assistant survey (Lee et al., 2011) | setting: nursing home. |
| 73 | 采取护士与护工捆绑管理的老年护理模式家属满意度调查（王丽 et al., 2016); Nurses and care workers bundled management aged care model: family member satisfaction survey, Wang L, Wang J, & Cong Y, 2016. | setting: a gerontological ward. |
| 74 | †分层次管理模式在护工全程服务中的研究（朱茂芳, 2012); Research on hierarchical management model in the whole process service of nursing attending workers, Zhu M, 2012. | participant: demographics of NAs was not performed. |
| 75 | †分层次护理模式下规范化培训对护理人员自我效能与职业倦怠的影响（谢玉兰 et al., 2013); Effect of standardized training on paramedics' self-efficacy and job burnout under hierarchical nursing model, Xie Y, Cao, J, & Guo S, 2013. | participant: all paramedic staff, and NAs were not discussed. |
| 76 | †分权管理模式在骨科护工服务管理中的应用（王晓琼 et al., 2021); Application of decentralized management model in service management of orthopaedic nursing attending workers, Wang X, Ge G, Shi J, & Shen T, 2021. | methodology: evaluation methods were not presented. |
| 77 | 管床护士与护工捆绑管理的老年护理服务模式探索（李雪, 2014); Generation of a tie-up between nurses and carers for geriatric nursing care, Li X, 2014. | specific area: the conclusion was only for geriatric nursing. |
| 78 | †护工层级管理对老年患者服务质量影响的探讨（陈萍 & 梁健, 2019); Exploration on effect of hierarchical management of nursing attending workers on service quality for the elderly, Chen P, & Liang J, 2019. | methodology: the intervention was mixed and blurred. |
| 79 | †护工统一管理模式在我院护理管理中实施的效果分析（李娟玲, 2016); Effect analysis of unified management mode of nursing attending workers in nursing management of our hospital, Li J, 2016. | methodology: evaluation tools (NAW competency test) were not presented detailly. |
| 80 | †护工星级管理模式用于外科患者的价值（沈玉美 et al., 2019); Value of the Star Management Model of nursing attending workers for surgical patients, Shen Y, Zhang X, & Xing X, 2019 | methodology: evaluation tools (NAW satisfaction questionnaire) were not presented detailly. |
| 81 | †护工星级管理模式在医院无陪护理管理中的应用效果探究（古晓莉 et al., 2020); Effect of nursing attending worker Star Management Mode in hospital unaccompanied nursing management, Gu X, Huang K, & Liu L, 2020. | methodology: evaluation tools (questionnaires on care quality and satisfaction of NAWs) were not presented detailly. |
| 82 | †护工主管负责制在神经外科护工管理中的应用（杨晓兰 et al., 2017); Application of the supervisor responsibility system in the management of nursing attending workers in neurosurgery, Yang X, Wan C, Zhong F, & Gui S, 2017. | methodology: evaluation tools (a satisfaction questionnaire and an NAW care quality questionnaire) were not presented detailly. |
| 83 | †护工自行管理在住院肿瘤患者中的应用价值研究（何应英 & 越太迁, 2014); Study on the application value of self-management of nursing attending workers in hospitalized cancer patients, He Y, & Yue T, 2014. | methodology: evaluation tools (satisfaction questionnaires for patients and their family members) were not presented detailly. |
| 84 | 护理员递进循环式与传统式教育培训效果的探索（徐筠, 2012); Nursing member of progressive exploration of the cycle and traditional education and training effect, Xu Y, 2012. | setting: nursing home |
| 85 | 护理员规范化管理中多元化策略的实践效果（袁赛霞 & 史定妹, 2013); Impact of the diverse strategy applying on the nursing assistant standardized management, Yuan S, & Shi D, 2013. | methodology: detailed evaluation methods were not performed. |
| 86 | †护理员团队化管理在老年住院患者生活照护中的应用（张玉玲 et al., 2018); Application of nursing assistants' team management in life care of elderly inpatients, Zhang Y, Ma Q, & Pu H, 2018. | methodology: evaluation tools (satisfaction questionnaire for patients) were not presented detailly. |
| 87 | †护士与陪护公司协同管理护工的老年护理服务模式探索（田志禾 et al., 2019); Exploration on elderly nursing service mode of nursing attending workers managed by nurses and escort companies, Tian Z et al., 2019. | methodology: evaluation tools (satisfactory of patients, working attitude of NAWs, and ward environment) were not presented detailly. |
| 88 | 基于 7S 下医护工一体化管理对重症医学科"三管"感染控制效果的对比分析（李萍 et al., 2020); Comparative analysis of the effect of "three management" infection control in ICU based on the integrated management of medical and nursing workers under 7S, Li P, Wang B, & Zhang Z, 2020. | participants were mixed (all staff in ICU) and NAs were not analyzed separatedly. |
| 89 | 基于岗位职责的三级护工管理模式在优质护理病房的应用（何琳 et al., 2020); The application experience of three-level nursing worker management model based on position responsibility in high-quality care, He L, Chen F, Zhou P, & Li H, 2020. | methodology: evaluation tools (care quality assessment and satisfactory of patients and doctors) were not presented detailly. |
| 90 | †金字塔式管理模式在无陪护工作中的应用体会（张惠贤 et al., 2011); Application of the pyramid management mode in unaccompanied care work, Zhang H et al., 2011. | type: this article was an introduction to a management mode and was not a research paper. |
| 91 | †老年干部病房中管床护士与护工信息化捆绑管理模式的建立与成效（贾倩, 2021); Establishment and effect of information binding management mode between bed nurses and nursing attending workers in elderly cadre wards, Jia Q, 2021. | methodology: intervention was not performed detailly. |
| 92 | †临床后勤服务部护工考评体系效果评价（冯梅 & 丁福, 2015); Effect evaluation of nursing worker evaluation system in clinical Logistics Service Department, Feng M, & Ding F, 2015. | methodology: evaluation methods were not performed detailly. |
| 93 | †某医院老年科护工绩效考核的研究与实践（董燕 et al., 2013a); Research and Practice on performance appraisal of nursing attending workers in geriatrics department of a hospital, Dong Y, Wang J, & Yang Y, 2013. | methodology: evaluation tools (for patients and family members) were not presented detailly. |
| 94 | 脑血管病患者医养结合模式中护理员教育培训体系的建立（朱红燕, 2016); The establishment on educational training system for caregiver in medical support binding models for patients with cerebrovascular disease, Zhu H, 2016. | setting: a nursing home-like facility. |
| 95 | †品管圈在提高干部病房护工管理质量中的应用研究（章芳芳 et al., 2017); Application of quality control circle in improving the management quality of nursing attending workers in cadre wards, Zhang F et al., 2017. | methodology: outcome measurement was not performed. |
| 96 | †情景模拟培训降低护理员照护老年患者跌倒发生率的应用研究（郭妍 & 邬小荣, 2020); Application of scenario simulation training to reduce the incidence of falls in nursing assistants care of elderly patients, Guo Y, & Wu X, 2020. | setting and specific area: this research was conducted in a geriatric hospital, and was only for fall prevention. |
| 97 | †实施优质护理服务 转变护工管理模式（陈晓欢 et al., 2013); Implementing high-quality nursing service and changing nursing attending worker management mode, Chen X, Song N, & He P, 2013. | methodology: evaluation methods were not presented. |
| 98 | †舒适护理理论在护工培训中的运用（陈华燕, 2015); Application of comfort nursing theory in nursing attending worker training, Chen H, 2015. | methodology and outcome: intervention method was not presented detailly, and there were not detailed outcomes. |
| 99 | †四位一体模式在优质护理服务病区管理中的应用（覃伟英, 2012); Application of "four in one" model in ward management of high-quality nursing service, Qin W, 2012. | participant: this article did not focus on NAs or NAWs. |
| 100 | 体验式教学在失能老人护理员培训中的应用研究（朱雅萍 et al., 2014); The experience of teaching in the application of the elderly nursing staff training to lose, Zhu Y, Wang J, & Xu T, 2014. | participant and setting: the participants were mixed (RNs, NAWs, and home carers) and NAWs were not discussed separately; and this study was conducted in a community care center. |
| 101 | †无陪护病房管理模式在消化内科的实施（郭丽艳, 2015); Implementation of unaccompanied ward management model in gastroenterology department, Guo L, 2015. | outcome: this article did not present detailed outcomes. |
| 102 | 无陪护模式病房护理人员能级管理体系的研究（杨雪莹, 2012); Study on competency-based grading system of clinical nurses in unattended wards, Yang X, 2012. | participant: this thesis was for paramedic staff, and NAs or NAWs were not included. |
| 103 | 无陪护医院医疗护理员培训体验的质性研究（高畅 et al., 2021); Qualitative research on the training experience of nursing assistants in unaccompanied hospitals, Gao C, Hao X, Song G, and Guo Y, 2021. | main focus: not on NA administration approaches. |
| 104 | 医院对精神科病区护工管理模式的探索与实践（乔辉英, 2016); Exploration and practice of the management mode of the nursing workers in the psychiatric department ward of the hospital, Qiao H, 2016. | main focus and specific area: this paper did not focus on NA administration approaches, and the conclusion was only fit for psychiatric wards. |
| 105 | 医院护工和保洁人员手卫生知识调查与培训（李荣明 et al., 2012); Hand hygiene knowledge investigation and training among assistant nurses and cleaning staff in hospitals, Li R, Jiang S, Chen X, & Luo B, 2012. | methodology and mixed participants: no detailed evaluation methods, and participants were mixed (NAs |

**Supplemental File 4** Reason for exclusion ($n$ = 112)

| No. | Publication | Excluded reason |
|---|---|---|
| | | and housekeepers) where NAs were not discussed separately. |
| 106 | †医院护理部与家政公司双重管理模式在护工管理中的应用效果（余立平 et al., 2019); Application effect of dual management mode of hospital nursing department and housekeeping company in nursing attending worker management, Yu L, Zeng L, & Guo Y, 2019. | methodology: evaluation tools (satisfactory of patients and healthcare staff) were not presented detailly. |
| 107 | †医院化管理在护工管理中的应用效果观察（代敏 & 黄晓红, 2017); Observation on the application effect of hospital management in nursing attending worker management, Dai M, & Huang X, 2017. | methodology: intervention methods were not presented detailly. |
| 108 | †优质护理病房护理员规范化管理分析（李霞, 2014); Analysis on standardized management of nurses in high quality nursing ward, Li X, 2014. | methodology: evaluation tools were not presented. |
| 109 | † "知-信-行"模式在神经外科护工培训的应用探讨（王爱丽 et al., 2013); Application of "knowledge-belief-practice" model in neurosurgical nursing worker training, Wang A, Xiong L, Zou Q, & Chen Y, 2013. | outcome: no outcomes. |
| 110 | †Barthel 指数评定量表在老年科护理员绩效考核的探索与实践（奚艳 et al., 2021); Exploration and practice of Barthel index evaluation scale in performance evaluation of nursing assistants in geriatrics department | methodology: evaluation tools (care quality and patient satisfactory) were not presented detailly. |
| 111 | **面向外来务工人员开展护理员技能培训与考核（邱翠琼 et al., 2017); Skill training and assessment of care worker for migrant workers, Qiu C, Wang N, Zhong Y, Pan S, & Tang X, 2017. | type: literature review. |
| 112 | ** †某医院老年科护工绩效考核的研究与实践（董燕 et al., 2013b); Research and Practice on performance appraisal of nursing attending workers in geriatrics department of a hospital, Dong Y, Wang J, & Yang Y, 2013. | methodology: evaluation tools (care quality) were not presented detailly. |

NA, nursing assistant / nursing aide; NNHS, National Nursing Home Survey; NNAS, National Nursing Assistant Survey; RN, registered nurse; HCA, healthcare assistant; NAW, nursing attending worker.

*: these articles did not have an abstract, so the abstract screening was not performed, and they were directly assessed for eligibility. ($n$ = 12)

**: these articles were manually retrieved from reference lists of included studies. ($n$ = 9)

†: titles and authors of these Chinese language articles were not from the original paper, and were translated into English by the authors themselves.
