## Supplementary material for "Administration approaches of nursing assistants in hospitals: a scoping review": online supplemental file 5

**Supplemental File 5** Fundings and conflicts of interest of included studies

| Study | Fundings | Conflicts of interest |
|---|---|---|
| Appleby 2019 | not reported | not reported |
| Campbell 2021a | not reported | no conflict of interest |
| Campbell 2020 | not reported | not reported |
| Dutton 2020 | not reported | not reported |
| Dykes 2011 | the Brigham and Women's Hospital Lily Kravitz Nursing Studies Award | no conflict of interest |
| Feng 2013 | funding from the Topics commissioned by the talent exchange service center of the Ministry of health | not reported |
| Friesen 2019 | not reported | not reported |
| Gao 2017 | not reported | not reported |
| Geoffrion 2020 | not reported | not reported |
| Haigh 2019 | not reported | no conflict of interest |
| Haraldsson 2021 | funding from Region Jönköping County, Sweden | no conflict of interest |
| Kennerly 2012 | the National Institute for Occupational Safety and Health Pilot Research Project Training Program of the University of Cincinnati Education and Research Center Grant No. T42/)H008432-06; a grant from the Robert Wood Johnson Foundation's Interdisciplinary Nursing Quality Research Initiative (INQRI) program; and the College of Nursing, University of Cincinnati, Dean's Award | no conflict of interest |
| Lee 2018 | not reported | not reported |
| Liu 2017 | not reported | not reported |
| Ma 2019 | not reported | not reported |
| McKenzie 2013 | not reported | not reported |
| Monteiro 2011 | not reported | not reported |
| Nie 2017 | not reported | not reported |
| Nørgaard 2012 | not reported | no conflict of interest |
| Pfeifer 2018 | not reported | no conflict of interest |
| Prestia 2012 | not reported | no conflict of interest |
| Qiu 2020 | not reported | no conflict of interest |
| Ritchie 2017 | grants application to the Guy's and St. Thomas' Charity S100414 | no conflict of interest |
| Small 2012 | not reported | not reported |
| Swann 2018 | not reported | not reported |
| Tom 2016 | not reported | not reported |
| Twigg 2016 | fundings from the Australian Research Council Linkage Projects, Sir Charles Gairdner Hospital and the Nursing and Midwifery Office, Department of Health Western Australia | no conflict of interest |
| Wagner 2018 | not reported | no conflict of interest |
| Ward 2014 | not reported | no conflict of interest |
| Wilson 2011 | grant by the Beaumont Foundation, Royal Oak, MI | not reported |
| Wu 2015 | not reported | not reported |
| Yu 2015 | not reported | not reported |
| Zhao 2020 | not reported | no conflict of interest |
| Zhi 2021 | supported by the National Health Commission of China [National Healthcare Improvement Initiative 2019] | no conflict of interest |
| Zhu D. 2021 | not reported | not reported |
| Zhu F. 2019 | not reported | not reported |
