## Supplementary material for "Administration approaches of nursing assistants in hospitals: a scoping review": online supplemental file 6

**Supplemental File 6** Quality appraisal outcome of included studies

S6.1 Overall scores

| Study | Tools | Grade |
|---|---|---|
| Appleby 2019 | MMAT | 80% |
| Campbell 2021a | MMAT | 100% |
| Campbell 2020 | AMSTAR 2 | Very Low |
| Dutton 2020 | MMAT | 60% |
| Dykes 2011 | MMAT | 100% |
| Feng 2013 | MMAT | 60% |
| Friesen 2019 | AMSTAR 2 | Very Low |
| Gao 2017 | MMAT | 40% |
| Geoffrion 2020 | AMSTAR 2 | High |
| Haigh 2019 | MMAT | 60% |
| Haraldsson 2021 | MMAT | 60% |
| Kennerly 2012 | MMAT | 80% |
| Lee 2018 | MMAT | 40% |
| Liu 2017 | MMAT | 60% |
| Ma 2019 | MMAT | 60% |
| McKenzie 2013 | MMAT | 60% |
| Monteiro 2011 | MMAT | 60% |
| Nie 2017 | MMAT | 80% |
| Nørgaard 2012 | MMAT | 60% |
| Pfeifer 2018 | MMAT | 40% |
| Prestia 2012 | MMAT | 60% |
| Qiu 2020 | MMAT | 80% |
| Ritchie 2017 | MMAT | 40% |
| Small 2012 | MMAT | 60% |
| Swann 2018 | MMAT | 80% |
| Tom 2016 | MMAT | 80% |
| Twigg 2016 | MMAT | 60% |
| Wagner 2018 | MMAT | 80% |
| Ward 2014 | MMAT | 80% |
| Wilson 2011 | MMAT | 60% |
| Wu 2015 | MMAT | 60% |
| Yu 2015 | MMAT | 60% |
| Zhao 2020 | MMAT | 60% |
| Zhi 2021 | MMAT | 60% |
| Zhu D. 2021 | MMAT | 60% |
| Zhu F. 2019 | MMAT | 60% |

S6.2 Original studies (using MMAT)

| Study | S1 | S2 | Type | Q1 | Q2 | Q3 | Q4 | Q5 |
|---|---|---|---|---|---|---|---|---|
| Monteiro 2011 | Y | Y | 4 | ? | Y | Y | Y | Y |
| Appleby 2019 | Y | Y | 5 | Y | Y | Y | ? | Y |
| Campbell 2021a | Y | Y | 4 | Y | Y | Y | Y | Y |
| Dutton 2020 | Y | Y | 3 | ? | Y | Y | N | Y |
| Dykes 2011 | Y | Y | 5 | Y | Y | Y | Y | Y |
| Feng 2013 | Y | Y | 4 | ? | Y | Y | Y | ? |
| Gao 2017 | Y | Y | 3 | ? | Y | N | N | Y |
| Haigh 2019 | Y | Y | 5 | Y | Y | Y | N | ? |
| Haraldsson 2021 | Y | Y | 4 | ? | Y | Y | ? | Y |
| Kennerly 2012 | Y | Y | 4 | Y | Y | Y | N | Y |
| Lee 2018 | Y | Y | 5 | Y | Y | ? | N | ? |
| Liu 2017 | Y | Y | 3 | ? | Y | Y | N | Y |
| Ma 2019 | Y | Y | 3 | ? | Y | Y | N | Y |
| McKenzie 2013 | Y | Y | 1 | ? | Y | Y | ? | Y |
| Nie 2017 | Y | Y | 4 | N | ? | Y | Y | Y |
| Nørgaard 2012 | Y | Y | 5 | Y | Y | ? | Y | Y |
| Pfeifer 2018 | Y | Y | 5 | Y | Y | Y | ? | N |
| Prestia 2012 | Y | Y | 3 | Y | ? | N | N | Y |
| Qiu 2020 | Y | Y | 3 | ? | Y | Y | N | Y |
| Ritchie 2017 | Y | Y | 3 | Y | Y | Y | N | Y |
| Small 2012 | Y | Y | 5 | Y | ? | Y | N | N |
| Swann 2018 | Y | Y | 3 | Y | Y | Y | ? | N |
| Tom 2016 | Y | Y | 3 | Y | Y | Y | N | Y |
| Twigg 2016 | Y | Y | 3 | Y | Y | Y | N | Y |
| Wagner 2018 | Y | Y | 3 | N | Y | Y | N | Y |
| Ward 2014 | Y | Y | 3 | Y | Y | Y | N | Y |
| Wilson 2011 | Y | Y | 3 | Y | Y | Y | ? | Y |
| Wu 2015 | Y | Y | 3 | ? | Y | Y | N | Y |
| Yu 2015 | Y | Y | 3 | N | Y | Y | N | Y |
| Zhao 2020 | Y | Y | 3 | ? | Y | Y | N | Y |
| Zhi 2021 | Y | Y | 4 | ? | Y | Y | N | Y |
| Zhu D. 2021 | Y | Y | 4 | N | ? | Y | Y | Y |
| Zhu F. 2019 | Y | Y | 3 | ? | Y | Y | N | Y |

Type: 1=qualitative study, 2=randomized quantitative study, 3=non-randomized quantitative study, 4=descriptive quantitative study, 5=mix methods study; Y=Yes, ?=can't tell, N=No

S6.3 Evidence-based reviews (using AMSTAR 2)

| AMSTAR 2 | | 1 | 2* | 3 | 4* | 5 | 6 | 7* | 8 | 9* | 10 | 11* | 12 | 13* | 14 | 15* | 16 | Overall Confidence |
|---|---|---|---|---|---|---|---|---|---|---|---|---|---|---|---|---|---|---|
| No. | Review | PICO | Protocol | Type Selection | Search | Duplicate Selection | Duplicate Extraction | Exclusion | Characteristics | RoB Assesment | Funding of Study | Appropriate Method | Impact of RoB in Synthesis | RoB in Discussion | Heterogeneity | Publication Bias | Conflict of Interest | |
| 1 | Campbell 2020 | Y | N | Y | P | Y | N | N | Y | N/A | N | N/A | N/A | N/A | N/A | N/A | N | Very Low |
| 2 | Frisen 2019 | Y | N | N | P | N | N | N | Y | N/A | N | N/A | N/A | N/A | N/A | N/A | N | Very Low |
| 3 | Geoffrion 2020 | Y | Y | Y | Y | Y | Y | Y | Y | Y | Y | Y | Y | Y | Y | Y | N | High |

Y=Yes, P=Partial Yes, N=No, N/A=not applicable
