## Supplementary material for "Administration approaches of nursing assistants in hospitals: a scoping review": online supplemental file 7

**Supplemental File 7** The frequency effect size of the topics and educational strategies

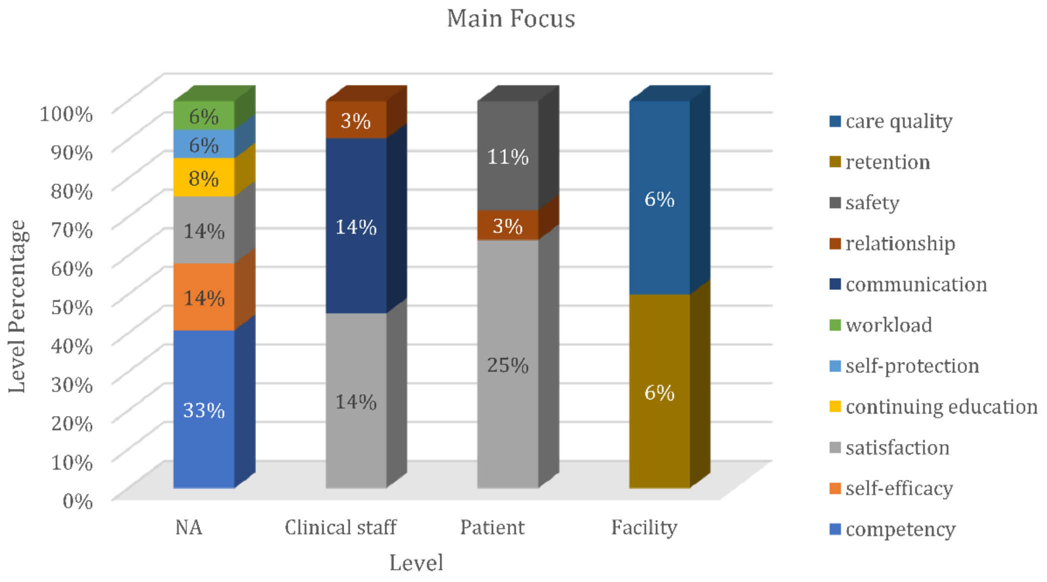


(a)

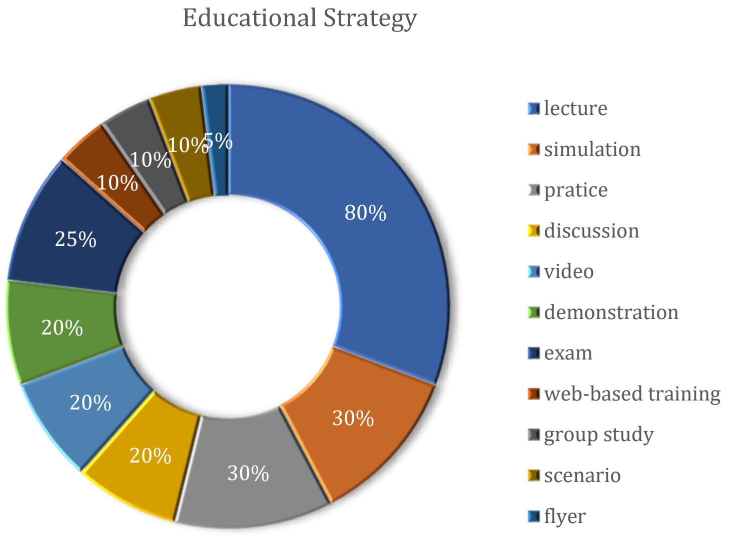


(b)

**Figure S1** The frequency effect size of the topics and educational strategies

(a) Main focus; (b) Educational strategy
